# A scalable serology solution for profiling humoral immune responses to SARS-CoV-2 infection and vaccination

**DOI:** 10.1101/2021.10.25.21265476

**Authors:** Karen Colwill, Yannick Galipeau, Matthew Stuible, Christian Gervais, Corey Arnold, Bhavisha Rathod, Kento T Abe, Jenny H Wang, Adrian Pasculescu, Mariam Maltseva, Lynda Rocheleau, Martin Pelchat, Mahya Fazel-Zarandi, Mariam Iskilova, Miriam Barrios-Rodiles, Linda Bennett, Kevin Yau, François Cholette, Christine Mesa, Angel X Li, Aimee Paterson, Michelle A Hladunewich, Pamela J Goodwin, Jeffrey L Wrana, Steven J Drews, Samira Mubareka, Allison J McGeer, John Kim, Marc-André Langlois, Anne-Claude Gingras, Yves Durocher

## Abstract

**OBJECTIVES:** Antibody testing against severe acute respiratory syndrome coronavirus 2 (SARS-CoV-2) has been instrumental in detecting previous exposures and analyzing vaccine-elicited immune responses. Here, we describe a scalable solution to detect and quantify SARS-CoV-2 antibodies, discriminate between natural infection- and vaccination-induced responses, and assess antibody-mediated inhibition of the spike-angiotensin converting enzyme 2 (ACE2) interaction.

**METHODS:** We developed methods and reagents to detect SARS-CoV-2 antibodies by enzyme-linked immunosorbent assay (ELISA). The main assays focus on the parallel detection of immunoglobulin (Ig)Gs against the spike trimer, its receptor binding domain (RBD), and nucleocapsid (N). We automated a surrogate neutralization (sn)ELISA that measures inhibition of ACE2-spike or -RBD interactions by antibodies. The assays were calibrated to a World Health Organization reference standard.

**RESULTS:** Our single-point IgG-based ELISAs accurately distinguished non-infected and infected individuals. For seroprevalence assessment (in a non-vaccinated cohort), classifying a sample as positive if antibodies were detected for ≥ 2 of the 3 antigens provided the highest specificity. In vaccinated cohorts, increases in anti-spike and -RBD (but not -N) antibodies are observed. We present detailed protocols for serum/plasma or dried blood spots analysis performed manually and on automated platforms. The snELISA can be performed automatically at single points, increasing its scalability.

**CONCLUSIONS:** Measuring antibodies to three viral antigens and identify neutralizing antibodies capable of disrupting spike-ACE2 interactions in high-throughput enables large-scale analyses of humoral immune responses to SARS-CoV-2 infection and vaccination. The reagents are available to enable scaling up of standardized serological assays, permitting inter-laboratory data comparison and aggregation.

## INTRODUCTION

The severe acute respiratory syndrome coronavirus 2 (SARS-CoV-2) pandemic has already induced four major waves of infection in Canada, resulting in > 3.2M confirmed infections and > 36K deaths (as of February 25^th^, 2022 ^1^). Overall seroprevalence estimates from natural infection during the pre-Omicron waves were relatively low compared to other countries ^2, 3^, and protecting the population from coronavirus disease of 2019 (COVID-19) has therefore heavily relied on vaccination. Fortunately, the dramatic acceleration of vaccination in Canada over the past 8 months has decreased symptomatic infections, severe disease, and deaths ^4^; as of February 18^th^, 2022, 84% of the population 5 years and older were fully vaccinated ^5^.

However, several important questions remain regarding both infection- and vaccination-induced humoral immunity, including the duration and decay of the immune response, the generation of functional neutralizing antibodies, and the overall differences in humoral responses across groups of individuals with different co-morbidities or following vaccination with different brands and regimens. This information can help guide and prioritize public health programs, such as vaccine booster schedules ^6^.

Plate-based enzyme-linked immunosorbent assays (ELISAs) are widely used scalable methods to quantitatively assess antibody responses to pathogens, including SARS-CoV-2. Microwells are coated with recombinant SARS-CoV-2 protein antigens, and biofluid samples (e.g. serum, plasma, dried blood spot (DBS) eluate, or saliva) are added (Figure 1a). If antibodies that recognize the antigen are present, they are detected with a secondary antibody, typically linked to an enzyme (such as horseradish peroxidase (HRP)) that elicits a measurable change upon addition of a colorimetric or chemiluminescent substrate.

**Figure 1.**
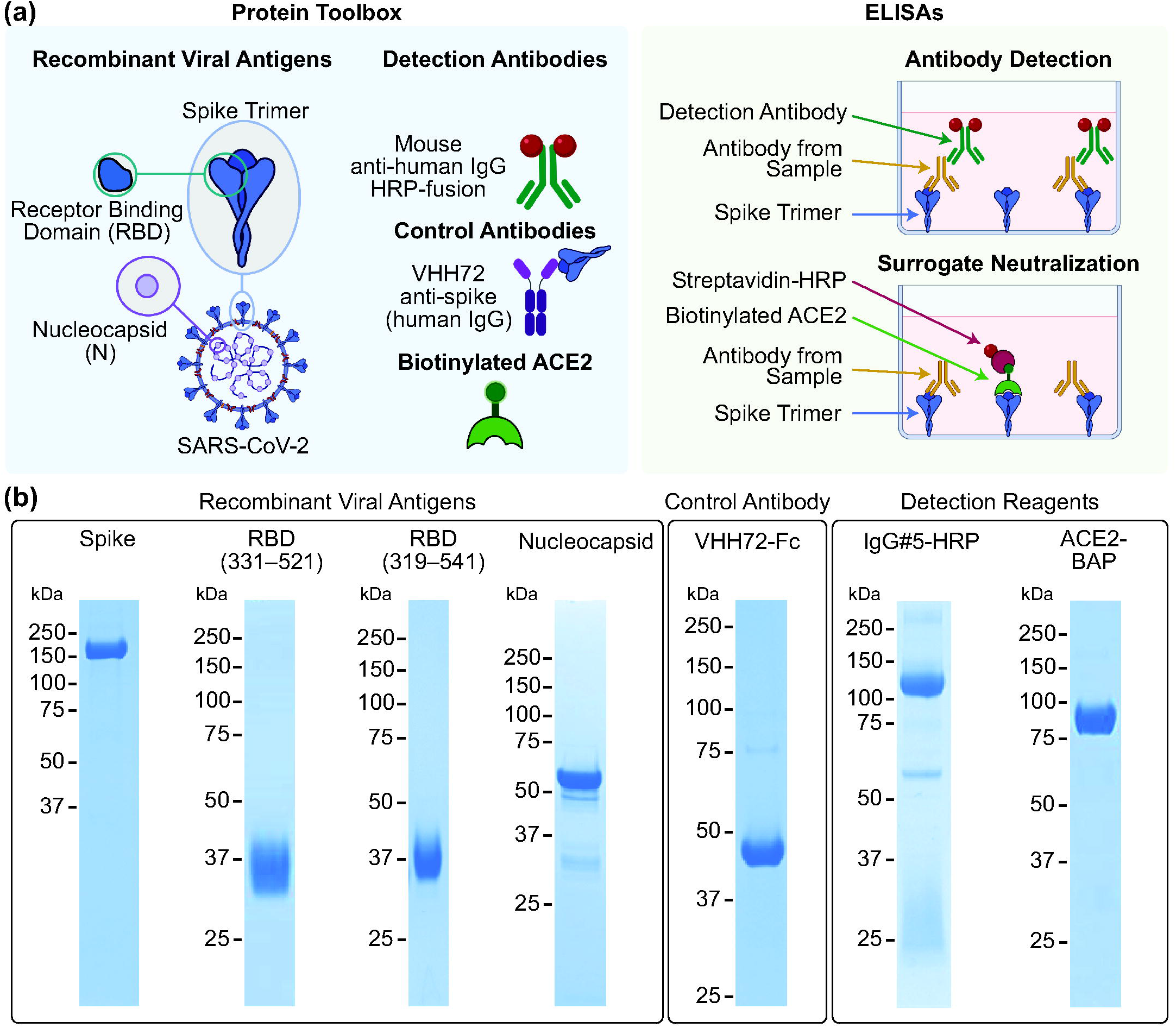
High-quality reagents for SARS-CoV-2 serology. **(a)** Reagents comprising the protein toolbox (left panel) are used in high-throughput plate-based ELISAs for antibody detection and surrogate neutralization (right panel). **(b)** The reagents were analyzed on Coomassie-stained polyacrylamide gels under reducing conditions to assess their purity. Molecular weight markers (kDa) are shown to the left of the gels.

Because of their potent immunogenicity, antigens derived from the SARS-CoV-2 spike and nucleocapsid (N) proteins have been most frequently used in serological assays, including those developed by commercial vendors. Full proteins, protein segments, and even peptides have been used in these assays which, combined with their different formats and readouts, yield distinct sensitivity and specificity profiles. This can lead to confusion in interpreting results. For example, conclusions about the persistence of antibodies to the nucleocapsid (N) protein differ depending on the platform used, and commercial vendors often do not disclose the exact amino acid sequence of their antigens ^7^.

We have previously used ELISAs to show that immunoglobulin (Ig)G (but not IgM or IgA) against SARS-CoV-2 spike, RBD, and N can persist for at least 3–4 months in the serum and saliva of individuals with COVID-19 ^8^, an observation now corroborated and extended by several other studies describing the persistence of circulating IgGs up to 13 months (although with gradual declines; e.g. ^9-12^). We also showed that the production of neutralizing antibodies capable of preventing interactions between spike (or its RBD) and its target angiotensin converting enzyme 2 (ACE2) could be monitored using a plate-based surrogate neutralization (sn)ELISA ^13^. We demonstrated good correlations between this assay and both spike-pseudotyped lentiviral and authentic SARS-CoV-2 plaque neutralization assays, suggesting that this could provide a scalable and suitable alternative to costly and labor-intensive classical antibody neutralization assessment.

While laboratory-based ELISAs like ours have been developed around the world, the various sources of antigens and antibodies used by different groups (in addition to batch-to-batch variation within groups) impede national and international comparisons of seroprevalence and vaccine effectiveness. This is further challenged by the lack of agreed-upon reference standards, with the exception of pools of convalescent plasma distributed through the World Health Organization (WHO) to calibrate assays to standardized units ^14^. Furthermore, assay performance variability over time remains incompletely examined. To address this, we have developed a reproducible and scalable SARS-CoV-2 serology solution using standardized protein reagents and protocols (Figure 1) and independent automated platforms in 2 Canadian laboratories in Toronto and Ottawa. This system has now been used to profile > 150K unique samples, demonstrating its scalability and robustness. Importantly, these reagents and protocols are available to the research community to enable implementation of these assays across laboratories.

## Results

### Reagent production

To facilitate comparison of SARS-CoV-2 serology results across Canada, we sought to develop and validate a scalable ELISA-based assay that leverages the mammalian protein expression expertise of the National Research Council of Canada (NRC) Human Health Therapeutics Research Centre. We optimized the production and purification of SARS-CoV-2 proteins (spike trimer, spike RBD, and N from the original Wuhan-Hu-1 strain) to generate large-scale high-purity batches of antigens (Figure 1b, Supplementary figures 1 and 2, Supplementary table 1). These were fused with purification tags ((His)_6_ and FLAG, with or without a Twin Strep-tag) and expressed in Chinese Hamster Ovary (CHO) cells either as stably transfected pools (spike) or through transient transfection (RBD, N) ^8, 15^. Antigens were purified by Immobilized Metal Affinity Chromatography (IMAC), with one or two additional purification steps performed for RBD and N (StrepTrap XT columns and/or preparative size exclusion chromatography, SEC). The spike and RBD proteins were highly pure when analyzed by SDS-PAGE (Figure 1b). By analytical SEC, spike eluted as a major (>95% integrated area) symmetrical peak of 490 kDa (consistent with its trimeric structure) with < 3% hexamers/aggregates, and both versions of RBD (encoding amino acids 319–541 and 331–521, respectively) eluted as single peaks with >95% integrated area. For purified N, some truncated forms were visible by SDS-PAGE/Coomassie staining; however, by SEC, it eluted as a major (>99% integrated area) peak with no apparent larger aggregates. Estimation of molecular weight of N was not possible by multiangle light scattering (MALS), but based on the SEC elution volume, it appears to be ∼300 kDa. This is consistent with the formation of tetramers or hexamers by recombinant coronavirus N, as reported by others ^16^. For recombinant protein production in CHO cells, post-purification yields ranged from 25 mg L^-1^ for RBD 331–521 to 370 mg L^-1^ for spike (Supplementary table 1).

The antigen toolbox was supplemented with a detection reagent, namely an anti-human IgG monoclonal antibody (IgG#5) expressed as a HRP fusion in-frame to the heavy chain (HC) C-terminus. This reagent was developed to address an issue common to HRP-based detection reagents from commercial sources, which are usually polyclonal anti-IgG antibodies conjugated to HRP post-purification. These commercial reagents display variability due to the nature of the polyclonal serum used by each supplier, as well as the ratio of conjugated HRP molecules per antibody. IgG#5 was produced by co-transfecting HC and light chain (LC) expression vectors in CHO cells. As shown in Figure 1, the purified antibody consists of a predominant band at ∼110 kDa, which represents the full-length HC-HRP fusion peptide. A minor band at ∼55 kDa is likely a form of the HC lacking HRP (confirmed by western blotting with an anti-mouse-Fc antibody (data not shown)), while the LC presents as a smear between 25 and 37 kDa. Because of this heterogeneity, purified IgG#5-HRP gave a convoluted profile by analytical SEC (Supplementary figure 1).

To have a common reference that enables intra- and inter-lab comparisons of SARS-CoV-2 antibody responses, we also generated recombinant anti-spike antibodies. Compared to pools of serum or plasma from convalescent patients, these offer the advantage of being renewable, scalable, and defined in sequence. Three recombinant single-domain antibodies (VHHs) were selected: VHH72 is a SARS-CoV and SARS-CoV-2 cross-reactive antibody described in Wrapp et al. ^17^, and NRCoV2-04 and - 20 are SARS-CoV-2 spike-binding VHHs developed in-house at the NRC. All antibodies were produced in CHO cells and purified on a MabSelect SuRe column. VHH sequences were fused at the C-terminus to Fc1X7 (an antibody-dependent cellular cytotoxicity (ADCC)-attenuated variant of human Fc) to produce dimeric proteins upon secretion from CHO cells. VHH72-Fc eluted as a single symmetrical peak of 102 kDa with < 2% aggregates, NRCoV2-04-Fc eluted with a major peak of 67% integrated area and a peak of 90 kDa, and NRCoV2-20-Fc as a single peak of 100 kDa with < 2% aggregates (Supplementary figures 1 and 2; Supplementary table 1). The toolbox was further supplemented with a biotinylated ACE2 protein (used for snELISA), which was purified on both IMAC and Strep-Tactin XT Superflow columns and elutes from SEC as a single peak of 90 kDa (Supplementary figure 1).

While the goal is ultimately to have a fully NRC-sourced protein toolbox, the current set of reagents is complemented by a commercial anti-N antibody to quantify the anti-N response (see Methods). As described below, the assays can also be repurposed for IgA/M monitoring with commercial secondary antibodies and matching reference antibodies for calibration.

### Direct detection ELISA

We previously developed ELISAs in 96-well colorimetric or 384-well chemiluminescent formats to assess the persistence of antibody responses^8^. This first version relied on small-batch research-grade or commercial reagents that are not readily scalable for nationwide surveillance studies. Here, we have first benchmarked the NRC reagents described above, and defined the optimal antigen and secondary antibody amounts in a 384-well automated format (see Methods). The NRC protein toolkit was independently optimized on automated platforms in Toronto (a Thermo Fisher Scientific F7 platform) and Ottawa (a Hamilton system), leading to minor variations in the optimal amounts of antigens and detection reagents, in part due to differences in the detectors on each system (Supplementary table 2). While the platforms can be used to generate full titration curves from serum samples, single-point measurements more readily enable scaling up to thousands of samples and have been used in most large-scale studies to date.

#### Toronto platform

The optimal dilution for single-point ELISAs was established using a 10-point four-fold titration series starting with 0.25 μL positive control serum per well (a 1:40 dilution, which is the amount used previously ^8^; Supplementary figure 3). A prozone (hook) effect was noted at the 1:40 dilution for samples with higher antibody levels, suggesting that a lower concentration was desirable. We performed receiver operating characteristic (ROC) analysis on a set of 300 negative pre-COVID19 samples (collected prior to November 2019) and 211 PCR-confirmed convalescent patients (>14 d post-symptom onset) at three different concentrations (Supplementary figure 4), optimizing for an amount of serum that would work with all three antigens. The 0.0625 μL per well serum condition was selected for single-point ELISA based on a reduced hook effect on spike (compared to the highest amount) and better specificity and sensitivity characteristics on its RBD (compared to the lowest amount). A final ROC analysis combining data from a second replicate at 0.0625 μL per well at a false positive rate of < 1% revealed sensitivities of 94%, 92%, and 81% for spike trimer, RBD, and N respectively (Table 1, Supplementary figure 4b). Importantly, replicates from experiments conducted 8 weeks apart were highly correlated (ρ > 0.87 for all three antigens, Figure 2a), similar to our previous observations ^8^, giving us confidence in conducting large-scale cohort studies.

**Table 1.** ELISA performance statistics for plasma or serum IgG (Toronto)

|  | spike | RBD | N | ≥ 2 positive antigens |
| --- | --- | --- | --- | --- |
| **ROC analysis** |  |  |  |  |
| **AUC** | 0.978 | 0.974 | 0.964 | n/a |
| **Threshold (Cut Point)** | 0.195 | 0.073 | 0.349 | n/a |
| **FPR** | 0.009 | 0.009 | 0.009 | 0.000 |
| **TPR** | 0.944 | 0.921 | 0.811 | 0.931 |
| **3 SDs from the mean of the negative controls** |  |  |  |  |
| **Threshold** | 0.190 | 0.186 | 0.396 | n/a |
| **FPR** | 0.011 | 0.000 | 0.007 | 0.000 |
| **TPR** | 0.944 | 0.890 | 0.786 | 0.912 |
| **Samples used to assess performance <sup>†</sup>** |  |  |  |  |
| Sample cohort | Replicates | Number | Total |  |
| **Patients with COVID-19** | 1 | 211 | 392 |  |
|  | 2 | 181 |  |  |
| **Pre-COVID-19 (negative)** | 1 | 300 for N and spike, 296 for RBD | 560 |  |
|  | 2 | 260 for N and RBD, 258 for spike |  |  |

<sup>†</sup>Negative samples were excluded from the analysis for spike and RBD if one replicate was positive.

**Figure 2.**
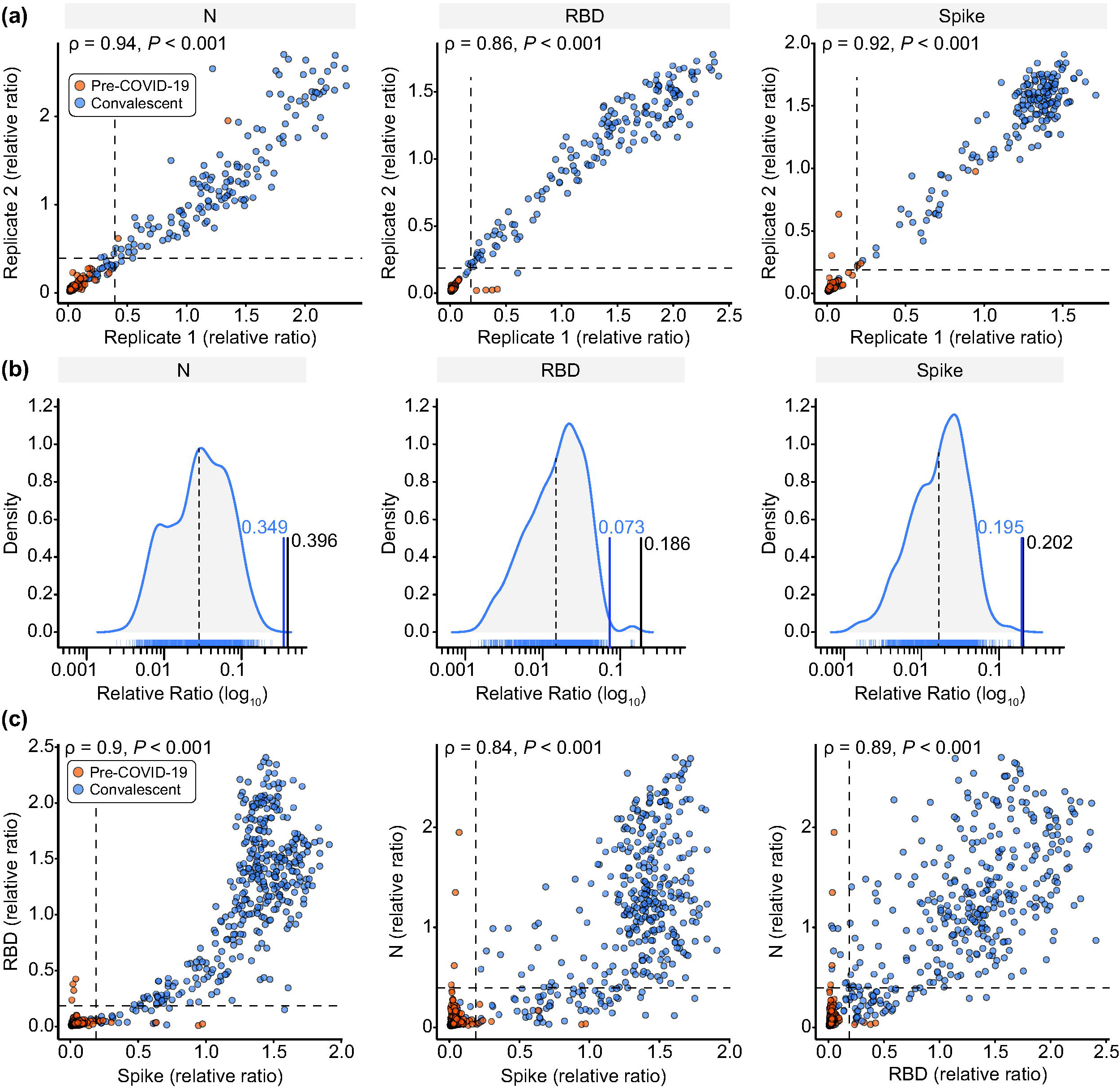
Development of high-throughput ELISAs for plasma or serum. **(a)** Known negative (pre-COVID-19) and positive (confirmed convalescent) samples (0.0625 µL/well) were tested in an automated antibody detection ELISA in two separate replicates 7 weeks apart. Spearman correlations are noted. **(b)** Density distributions of negative samples were plotted for each antigen. The black lines represent the mean of the negative distribution (dotted) and three SDs from the mean (solid; the relative ratio is indicated). The blue line represents the thresholds established by ROC analysis. **(c)** Comparison of the antigens with a set of known negative and positive samples at 0.0625 µL/well. Spearman correlations are shown. For both (a) and (c), dashed lines represent the thresholds as defined by the 3-SD negative distribution shown in (b) and listed in Table 1.

#### Ottawa platform

Early optimizations of the serological assay to identify the serum dilution within a linear range with the lowest non-specific reactivity to the negative serum were performed using the manual colorimetric ELISA (Supplementary figure 5). The colorimetric assay was used to test the performance of the HRP-conjugated antibodies, including six different anti-human IgG conjugate antibodies provided by the NRC (Supplementary figure 6a) with different characteristics and enzyme conjugation strategies (e.g. HRP fusion vs conjugation). While all candidates offered good detection, IgG#5—a direct HRP fusion—was selected for further assays, based on its superior quantitative capacity and extremely low non-specific reactivity within a standard reaction time (Supplementary figure 6b). This reagent was also compared to commercial polyclonal and monoclonal secondary antibodies for the detection of anti-spike, -RBD, and -N IgG in DBS samples (see below and Supplementary figure 7c). While all antibodies correctly identified negative and positive DBS samples, IgG-#5-HRP offered the greatest distinction between the negative and positive distributions. To maintain a high level of specificity, the secondary antibody concentration and the nature of substrate were modified between the manual and automated ELISAs (see Supplementary table 2 for the final concentrations).

### Setting positivity thresholds in seroprevalence studies

One concern with establishing thresholds based on a single sample set is that over time and with variations in sample type (venipuncture vs. capillary blood collection, different collection tubes and handling conditions, etc.), the background of the assay may change, affecting the threshold for reporting positives. In Toronto, we monitored the negative controls (blanks, pre-COVID-19 negative sera, commercially purified IgG, *n* = 1,320 for spike and 1,248 for N and RBD) used in 23 experiments conducted over 4 months. We calculated a threshold of 3 SDs from the mean of the log distribution of these controls and compared it to the thresholds established by ROC analysis (Figure 2b). The thresholds were very similar for N and spike, but the 3 SD range was more stringent for the RBD, decreasing the likelihood of false positive calls. We therefore adopted a threshold of 3 SDs from the mean of these controls for each antigen and recalculated our performance characteristics (Table 1). At this threshold, the specificity for RBD increases to 100% with a slight decrease in sensitivity to 89%, spike retained the same sensitivity and specificity (99% and 94%, respectively), whereas N’s sensitivity decreased from 81% to 79% with 99% specificity. The same strategy was applied to IgM and IgA to define stringent thresholds (Supplementary figure 8, Supplementary table 4).

Even with these conservative thresholds, an unacceptable number of false calls may be made in low seroprevalence situations (as in Canada, particularly during the first waves of infection). This is shown by the negative samples (red dots in Figure 2a) that passed the thresholds in individual assays. Importantly, these pre-COVID negatives were only ever positive in one test, in contrast to true positives, which tended to pass the positivity thresholds of ≥ 2 tests, consistent with the high correlation between the assays (ρ > 0.84 for all three combinations, with the highest correlation between spike and its RBD; Figure 2c). Our final determination of sample positivity in seroprevalence settings therefore requires that it exceeds the thresholds of at least 2 out of 3 antigens. As can be seen in Figure 2C, this strategy eliminates the false positive calls for the individual antigens (red dots) while retaining similar sensitivity (combined specificity of 100% and sensitivity of 91%).

### Harmonization between platforms and testing the WHO standards

To compare our ELISA-based assays with others, we tested them using WHO international reference panel 20/268, comprised of five samples (Negative, Low, Mid, High, and Low S/High N; Figure 3, Supplementary figure 9) and expressed the results in BAU mL^-1^ by comparing signals to the WHO International Standard (20/136). For this, we used single-point measurements from dilutions that were within the linear range of the International Standard curve (Toronto) or the median of three dilutions (Ottawa). For all five samples, the median results from both labs were highly correlated (ρ = 1 for spike and N, 0.9 for RBD) and were between 0.5–2 fold of the geometric mean (red line with half arrows) reported in a multi-lab comparison by WHO ^14^. Importantly, by applying a conversion formula to our relative ratios for expression in BAU mL^-1^ (Figure 3d, Supplementary Table 3), our results can be readily compared to other national or international efforts. Using this formula, plasma or serum seropositivity thresholds for IgG are 34, 31, and 11 BAU mL^-1^ for nucleocapsid, RBD, and spike respectively (Toronto). When samples are diluted 1:2560, the assays can detect values up to 5344, 4454 and 1500 BAU mL^-1^ for N, RBD, and spike respectively.

**Figure 3.**
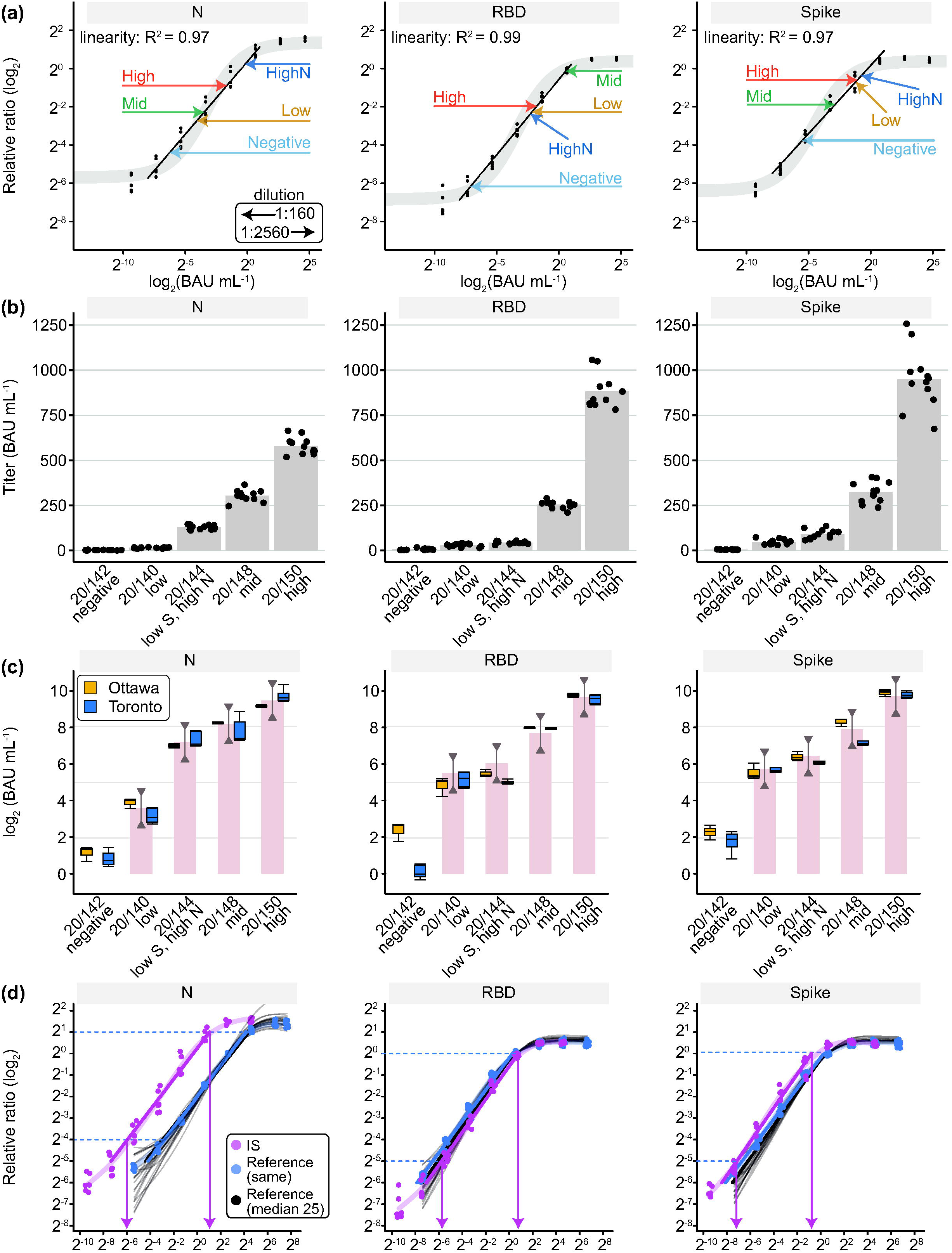
Conversion of Ottawa and Toronto ELISA data to WHO BAUs and comparison to the WHO Reference Panel 20/268. **(a)** The 20/268 reference panel at the indicated dilutions (arrows) was fitted onto a dose-response curve of the IS with the measured values expressed as relative ratios (Toronto). A 1:160 dilution (0.0625 µL/well) of sample was used except when it was out of the linear range of the fitted line in which case the 1:2560 dilution (0.0039 µL/well) was used. **(b)** IgG levels in the five samples in the 20/268 reference panel are represented for spike, its RBD, and N (n = 12, 4 replicates at 1:500, 1:1,000, 1:2,500 dilutions). **(c)** Box plots for Ottawa (orange) show the median of the 12 samples from B. Box plots for Toronto (blue) show the median of individual measurements (n = 4) for the selected dilution (1:2560 (0.0039 µL/well) for Mid and High for spike and N, High for RBD, the rest were at 1:160 (0.0625 µL/well)). The WHO bar graph shows the geometric mean from the WHO study and the lines with half arrows represent a 0.5–2-fold range from the geometric mean. **(d)** Reference curves (VHH72-Fc for spike/RBD, anti-N for N) were plotted for each antigen either from the same tests in which the IS was analyzed or from 25 different tests over 3 months (shown as faded black lines with a thicker median line in black). The blue dashed lines represent the limits of the linear intervals for the curves and the pink arrows represent the BAU mL^-1^ at those points. As the reference curves are parallel to the IS within the linear interval, a conversion factor can be applied to convert relative ratios to international BAU mL^-1^ units (Supplementary table 4). For illustrations purposes to show IS and reference curves in the same panel, the X-axis is BAU mL^-1^ for IS and μg mL^-1^ * 100 for the reference curves.

### DBS testing

For serosurveillance, it is important to capture broad swaths of the Canadian population, and at-home sample collection is ideal for this, especially during times of lockdown or restricted movement. DBSs are easier to collect, more stable at room temperature, and considered non-hazardous when dried ^18, 19^. We optimized the elution concentrations for both the colorimetric and automated assays (see Methods), and then compared the results of DBS samples and matching plasma samples using reference samples created by the National Microbiology Laboratory (NML; panel 1, with a total of 26 different DBSs and matching plasma samples; Supplementary figure 10a). Using NML panel 3, we also found a high correlation in signal between different punch sizes at a constant ratio of punch area to elution volume (Supplementary figure 10b).

However, despite this high correlation between the two sample types, the higher relative ratios of DBS samples compared to plasma or serum samples shifted the positive and negative density distributions, making direct application of the plasma/serum thresholds inappropriate for scoring seropositivity using DBSs (Supplementary figure 10c). ROC analysis was therefore performed in Toronto and in Ottawa on panel 4, also supplied by the NML (Tables 2 and 3, Supplementary figures 11 and 12), yielding sensitivities of 98% for spike and its RBD and 92% for N at a 1% false positive rate threshold in Toronto. By requiring 2 out of 3 calls to be positive, the specificity increased to 100% and the sensitivity was 98%. In Ottawa, at a 3% FDR, the assays achieved 100% sensitivity for each antigen with false positive rates of 2% for spike, 1% for its RBD, and 6% for N. When requiring 2 out of 3 calls to be positive, specificity and sensitivity were both 98%. In the final protocol, we opted to use two 3 mm (3.2 mm in Ottawa) punches, as this enables more flexibility and reproducibility when collections are not ideal (which would be typical of self-collection). The minimal sample requirement means the analysis can still be performed even when full DBSs are not provided.

**Table 2.** ROC statistics for the DBS IgG ELISA (Ottawa)

| Antigen | spike | RBD | N | ≥ 2 positive antigens |
| --- | --- | --- | --- | --- |
| AUC | 0.991 | 0.996 | 0.992 | N/A |
| Threshold (Cut Point) | 1.410 | 0.868 | 1.171 | N/A |
| FPR | 0.02 | 0.01 | 0.06 | 0.02 |
| TPR | 1.00 | 1.00 | 1.00 | 0.98 |
| **Number of samples used to generate ROC curves<sup>†</sup>** |  |  |  |  |
| Antigen | spike | RBD | N | ≥ 2 positive antigens |
| **Patients with COVID-19** | 97 | 97 | 97 | 97 |
| Number excluded | 2 | 2 | 2 | 2 |
| Total analyzed | 95 | 95 | 95 | 95 |
| **Pre-COVID-19 (negative)** | 90 | 90 | 90 | 90 |
| Number excluded | 0 | 0 | 0 | 0 |
| Total analyzed | 90 | 90 | 90 | 90 |

<sup>†</sup>NML panel 4. Each sample was analyzed once, and thresholds were defined at 3% FDR as described in Supplementary figure 11. Two positive samples were excluded as they were early on in infection.

**Table 3.** ROC statistics for the DBS IgG ELISA (Toronto)

| Antigen | spike | RBD | N | ≥ 2 positive antigens |
| --- | --- | --- | --- | --- |
| AUC | 0.99 | 0.99 | 0.99 | N/A |
| Threshold (Cut point) | 0.482 | 0.324 | 0.642 | N/A |
| FPR | 0.01 | 0.01 | 0.01 | 0.00 |
| TPR | 0.98 | 0.98 | 0.92 | 0.98 |
| **Number of samples used to generate ROC curves<sup>†</sup>** |  |  |  |  |
| Antigen | spike | RBD | N | ≥ 2 positive antigens |
| **Patients with COVID-19** | 97 | 97 | 97 | 97 |
| Number excluded | 0 | 0 | 0 | 0 |
| Total analyzed | 97 | 97 | 97 | 97 |
| **Pre-COVID-19 (negative)** | 90 | 90 | 90 | 90 |
| Number excluded | 3 | 1 | 2 | 4 |
| Total analyzed | 87 | 89 | 88 | 86 |

<sup>†</sup>NML panel 4. Each sample was analyzed in duplicate (the unique numbers of samples are shown). Replicates were treated as separate samples in the ROC analysis. Negative samples were excluded from the analysis if the replicates were > 4 SDs from the mean.

### snELISA

We previously reported an ELISA-based test that functions as a surrogate to more complex neutralization assays employing live viruses, at least with regards to assessing antibodies that prevent the interaction of spike with the ACE2 receptor ^13^. In this snELISA, a sample is added to a plate coated with RBD or spike antigens (Figure 1a). Instead of a secondary antibody, biotinylated ACE2 is added followed by streptavidin-HRP. If antibodies from the sample recognize the antigen, they will block biotinylated ACE2 from binding, leading to decreased signal when HRP substrate is added. The biotinylated ACE2 was first tested in a manual colorimetric assay using recombinant antibody and known negative and positive samples (Supplementary Figure 13). Both the full-length spike trimer and the RBD domain (319–541 construct) were comparable to prior reagents produced at smaller scale. The assay was then transferred to an automated 384-well chemiluminescent format. Using a panel of samples from the Canadian Blood Services and from two donors after one or two doses of the BNT162b2 (aka Comirnaty; Pfizer-BioNTech) mRNA vaccine, the dose response curves between spike and its RBD were significantly correlated (ρ = 0.90; Figure 4A, Supplementary figure 13).

**Figure 4.**
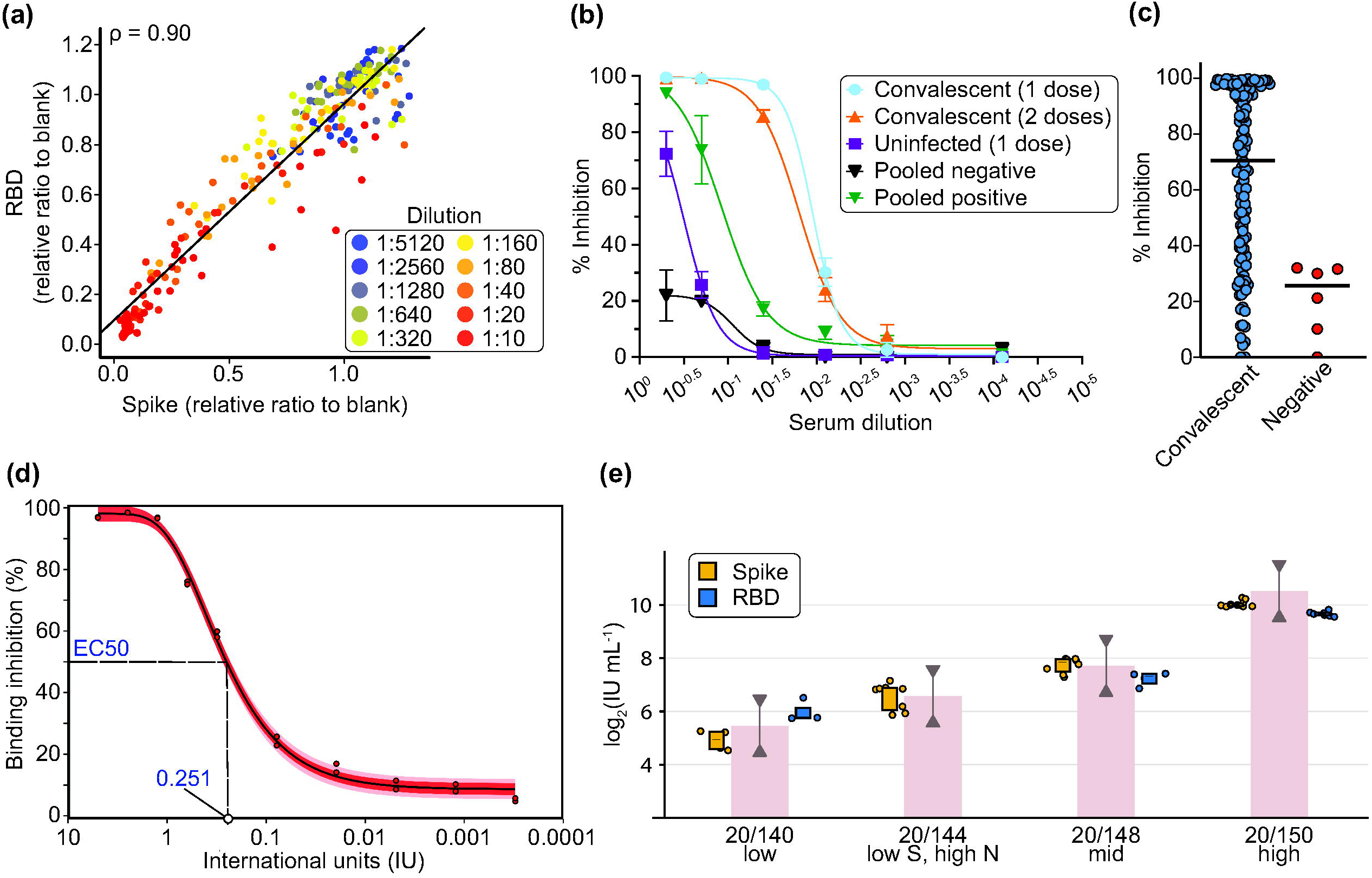
Dose response curves or single-point snELISA and conversion to International Units using the Who International Standard. **(a)** Correlation of spike to RBD as snELISA antigens is shown for 11 samples in a 10-point dilution series (individual curves are shown in Supplementary figure 13). **(b)** Dose response curves (*n* = 4) for the spike snELISA. Samples were from convalescent SARS-CoV-2 individuals 3 weeks after 1 or 2 doses of Comirnaty vaccine (Pfizer) or an uninfected individual (from a surveillance study) 3 weeks post-first dose of Comirnaty. Pooled sera were from 100 individuals with or without prior SARS-CoV-2 infection. **(c)** Single point measurements (at 1:5 dilution) using the spike snELISA. **(d)** Titration of the neutralizing activity of the WHO IS using snELISA. Raw luminescence values were converted to inhibition of ACE2-Spike binding; maximal signal (i.e. 0% inhibition) was measured in absence of convalescent plasma (PBS only). The normalized data was fitted with a four-parameter logistic function and the 95% confidence interval (IC95) and two standard deviations (2SD) is shown. **(e)** Box plots for the WHO reference panel 20/268 using RBD (blue, Toronto, ACE2 source: Rini) or spike (orange, Ottawa) as antigens. For RBD, *n* = 3 at 1:10 dilution for Low, *n* = 4 at 1:10 dilution for Mid, *n* = 7 at 1:40 and 1:160 for High. For spike, *n* = 12 for High (4 replicates at 3 dilutions), *n* = 8 for LowS HighN and Mid (4 replicates at 2 dilutions) and *n* = 4 for Low (2 replicates at 2 dilutions). The WHO bar graph shows the geometric mean from the WHO study and the lines with half arrows represent a 0.5–2-fold range from the geometric mean. No inhibition was seen for the LowS HighN sample for RBD.

The snELISA was also implemented on the Ottawa automated platform. Titration of the coating antigen, streptavidin-HRP polymer, and luminescent substrate were performed with varying amounts of biotinylated ACE2 to establish the concentrations with optimal sensitivity, appropriate dynamic range, and a minimized hook effect at high ACE2 concentrations (Supplementary figure 14). The final concentration of ACE2 per well was established at 6.5 ng, using 100 ng/well of coated antigen and the substrate diluted 1:2. Using these conditions, we were able to generate consistent neutralization efficiency measurements with serum and plasma (Figure 4B, Supplementary figure 15), enabling us to calculate effective concentration for 50% inhibition (EC50) values for convalescent sera and in sera from doubly vaccinated individuals. We also tested a panel of 121 positive and six negative samples from the *Stop the Spread Ottawa* study as single point measurements at a 1:5 dilution using spike as the antigen (Figure 4C). Convalescent samples ranged from 0–100% inhibition, with a median value of 70%. The six negative samples ranged from 0–32% inhibition with a median of 26%, consistent with our previous observation that pre-pandemic samples can achieve partial inhibition of the spike-ACE2 interaction due to cross-reactivity of antibodies targeting seasonal coronaviruses ^20^. For DBS samples, four 3 mm punches eluted in 100 μL of PBS from a double-vaccinated individual showed measurable neutralization activity (Supplementary figure 15); however, we were unable to measure neutralization in DBS samples from convalescent or singly vaccinated individuals (in contrast to serum). Therefore, while it is possible to detect neutralization activity from DBSs, this is limited to samples with high neutralizing activity. Additionally, DBS samples might not be ideal for quantitative neutralization measurements, as sample quality, paper saturation, and disc punching consistency can interfere with the reproducibility and reliability of the results.

The final development performed with the snELISA was to correlate binding inhibition to international units (IUs) of the WHO International Standard (Figure 4D). By titrating the standard, we established the correlation between binding inhibition and IUs within the linear range of the curve, with 0.251 IUs being required to inhibit ACE2-spike interaction by 50% (EC50). We then tested the WHO reference panel 20/268 by snELISA using spike (Ottawa) or RBD (Toronto) as antigens (Figure 3E). For the low, mid and high samples, results from Toronto and Ottawa were between 0.5–2 fold of the geometric mean (red line with half arrows) reported in a multi-lab comparison by the WHO ^14^. For the low S, high N sample, there was no inhibition of ACE2 binding in the snELISA using RBD as the antigen, in contrast to the snELISA using full-length spike which detected inhibition levels similar to the WHO study. Given the numerous different protocols and strategies that are being used to measure neutralizing antibodies such as spike-pseudotyped lentiviral assays, plaque reduction neutralization tests (PRNT) and the snELISA, transforming the data to IUs will enable more robust cross-laboratory and cross-assay correlations to be performed.

### Visualization of the results of seroprevalence and vaccination studies

As defined above, the assays optimally use the IgG responses to three antigens (spike, RBD, N) to report seropositivity resulting from SARS-CoV-2 infection. To illustrate how vaccination cohorts differ from infected cohorts, we re-plotted a subset of the data from our training set (positives and negatives) as pairwise comparisons of spike and N on scatterplots, with the color intensity mapping to that of RBD (Figure 5A). We also replotted data from a cohort of vaccinated patients on dialysis ^21^ prior to vaccination and 6 weeks afterward (Figure 5B). As expected, the infected cohort contained samples largely positive for all three antigens tested, while the vaccinated cohort largely consisted of samples positive for spike and its RBD, but negative for N, which is not targeted by vaccines used in Canada. This type of visualization helps define individuals with evidence of infection from those with antibodies resulting from vaccination.

**Figure 5.**
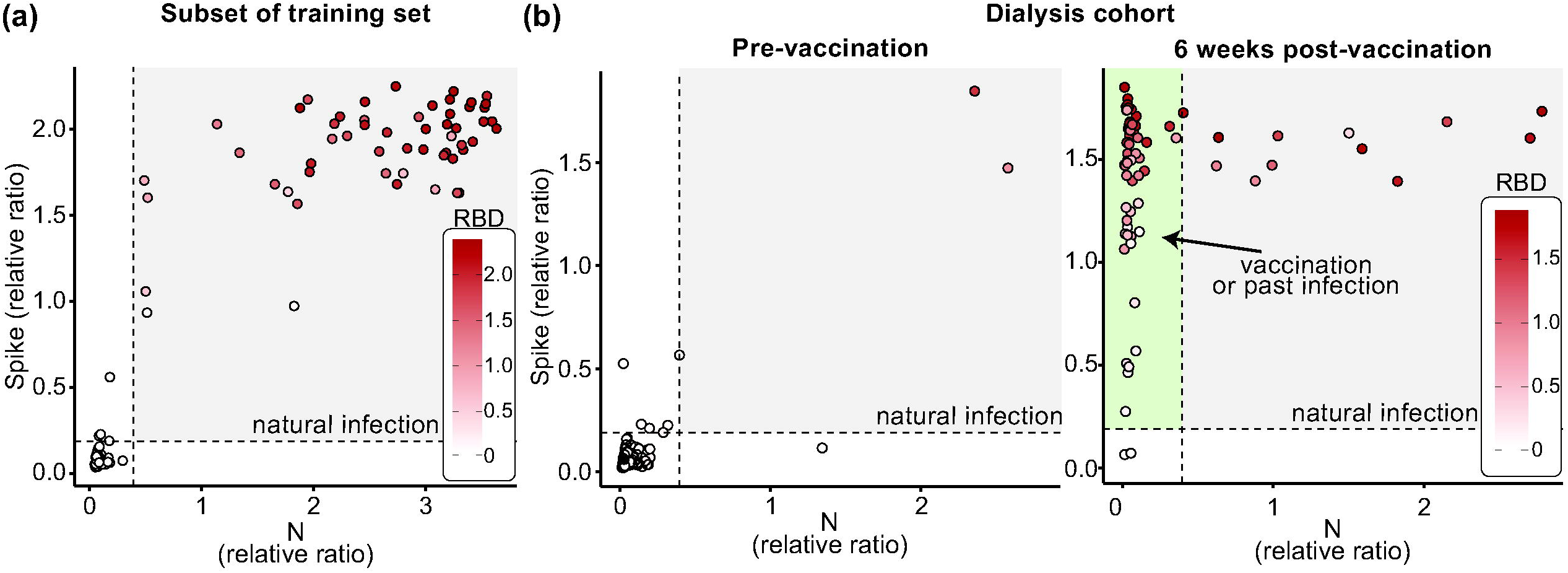
Visualization of Data from 3 Antigen testing. The results from the three antigens with known negative and positive samples **(a)** and samples from a longitudinal study of patients on dialysis at baseline and after their first vaccine dose **(b)**. The dashed lines represent the thresholds for spike and N. The area with positives for both spike and N (colored in gray) is indicative of natural infection and the area showing samples that are N-negative but spike-positive (and RBD-positive if colored) is highlighted in green on the right panel and is indicative of vaccination.

## Discussion

In this paper, we describe a toolkit and protocols for both SARS-CoV-2 serosurveillance and vaccination response profiling that we applied to populations studies in Canada. We have validated these reagents, produced and made available by the NRC, in two separate laboratories. The results can be expressed as relative ratios or can be converted into international units (BAU mL^-1^ for antibody detection, IU mL^-1^ for antibody neutralization) to facilitate comparisons between labs and with studies proposing correlates of protections based on these units ^22^. We validated antibody detection and surrogate neutralization assays in 96-well format (manual colorimetric assays) and on two different 384-well automated chemiluminescent platforms, enabling these reagents to be used at small-scale in laboratories with basic equipment and in dedicated high-throughput facilities equipped with different automation equipment.

All vaccines approved for use in Canada (also in USA and Europe) use spike as the immunogen ^23-25^. For those who have been vaccinated, responses to spike (and its RBD) are expected, whereas a response to N is only expected after infection. Thus, it is of value to focus on these two orthogonal readouts when detecting and characterizing immune responses ^26^. Adding the spike RBD as a third antigen increases specificity [of the three antigens, it is the least conserved in coronaviruses ^27-29^] and provides insight into possible protection, as it is also the antigen most correlated with neutralization potential ^13, 30, 31^.

By definition, single point assays must make compromises in defining antigen/antibody/detection antibody amounts to ensure that a large fraction of measurements are within the linear range of quantification. Here, we have optimized the concentrations of the serum/plasma/DBS eluates used to capture the most positives (i.e. convalescent individuals) possible without inducing a hook effect. Still, in samples with very high antibody levels (including those from healthy individuals following two doses of vaccine), the measured antibody levels will saturate the assay, thereby preventing their accurate measurement. When the relative amounts of antibodies detected across samples is important to measure, expanded dilutions should be performed (note that this drastically decreases assay throughput and increases costs). As a compromise, in vaccinated cohorts, we routinely perform two-point dilution (16-fold difference) assays that expand the range of concentrations that can be measured in the linear range of quantification, from 1 BAU mL^-1^ to 1500 BAU mL^-1^ for spike, and from 3 BAU mL^-1^ to 4454 BAU mL^-1^ for RBD.

We also optimized our seropositivity thresholds to limit false positives, both by providing a stringent definition of positivity in each assay, and by requiring that this positivity threshold be met in two separate assays to establish overall positivity in seroprevalence studies. While we felt this was essential to avoid inflating the positive calls in low prevalence settings, it may affect the overall sensitivity of the assays, especially following recent infection/vaccination (where seroconversion for anti-spike often precedes that of RBD and N ^8^) or as antibody levels decay. Other limitations stem from the selection of antigens, as those used in these assays are from the original Wuhan-Hu-1 strain, and it is possible that decreased sensitivity occurs when testing patients infected with variants, especially Omicron which has 30 amino acid substitutions (15 within the RBD), three small deletions and one small insertion in the spike coding sequence ^32^ [though our initial results with non-vaccinated Omicron-infected convalescents indicate detection of anti-RBD IgG on this assay; optimization of assays based on Omicron sequences is ongoing]. Lastly, as assays using N generally have poorer specificity and sensitivity (at our defined thresholds) than RBD- and spike-based assays, they may not be ideal as a stand-alone method to define vaccine breakthrough infections, unless serial blood samples are available (in which case an increase in anti-N levels would indicate that an infection has occurred). Further assay development will include additional viral antigens not contained in the vaccines to increase the detection sensitivity for antibodies produced in breakthrough infections. The control over reagent production and quality, and the flexibility of the ELISA platforms to accommodate new antigens also provide a rapid route to testing immune responses to variants. All reagents and protocols are publicly available to enable the rapid deployment of these assays.

The validated automated ELISA assays described here are currently being used in multiple Canadian studies (with >150,000 unique samples profiled to date). These include large serosurveys that monitor the global (and regional) humoral response (from both infection and vaccination) in the Canadian population, including the Canadian COVID-19 Antibody and Health Survey from Statistics Canada ^2^, seroprevalence studies with Canadian Blood Services ^33, 34^, the Action to Beat Coronavirus study ^35^, and the Canadian Partnership for Tomorrow’s Health study ^36^. Additionally, these assays are used in > 30 studies focused on infection and/or vaccine responses across different cohorts, e.g. in persons predicted to have a weaker immune response (based, for example on age ^37^ or health conditions ^38^), or who are likely to be exposed to infection from the workplace. The results of the assays have critically helped informed public health decisions. Based on, e.g. a weak response to vaccine in older adults or patients on dialysis ^21, 39^, or the decline in antibody levels post-dose 2 in residents of long-term care homes ^40, 41^, prioritizations were made for second doses, vaccine type was recommended, or additional doses were accelerated in the populations studied. Combined with other studies coordinated by the COVID-19 Immunity Task Force (CITF) ^42^, our serology studies help guide Canada’s response to the COVID-19 pandemic.

## Methods

### Protein production

#### Spike trimer

The SARS-CoV-2 spike ectodomain construct (SmT1) from the Wuhan-Hu-1 strain with S1/S2 furin site mutations, K986P/V987P prefusion-stabilizing mutations, and human resistin as a trimerization partner ^15^ was produced using stably transfected Chinese Hamster Ovary (CHO) pools (CHO^BRI/2353™^ cells) and purified as described ^8^. To prepare reference material aliquots for distribution by the National Research Council of Canada (NRC) Metrology Research Centre, the bulk purified protein was formulated in PBS supplemented with 10 mM HEPES sodium salt. After aliquoting and one freeze-thaw cycle, protein integrity and purity were assessed by sodium dodecyl sulfate-polyacrylamide gel electrophoresis (SDS-PAGE; Figure 1B) and analytical size-exclusion ultra-high performance liquid chromatography (SEC-UPLC; Supplementary figure 1, Supplementary table 1). SEC-UPLC was run on an Acquity H-Class Bio UPLC system (Wyatt Technology, Santa Barbara, CA, USA) in phosphate buffered saline (PBS) + 0.02% Tween-20 on a 4.6 × 300 mm Acquity BEH450 column (2.5 μm bead size; Waters Limited, Mississauga, ON, Canada) coupled to a miniDAWN Multi-Angle Light Scattering (MALS) detector and Optilab T-rEX refractometer (Wyatt).

#### Nucleocapsid

N cDNA (corresponding to amino acids 1–419 of YP_009724397, Wuhan-Hu-1 strain) was synthesized by GenScript (Piscataway, NJ, USA; using Cricetulus griseus codon bias) with a C-terminal FLAG-Twin-Strep-tag-(His)_6_ tag and cloned into the pTT5® expression plasmid (NRC) to create NCAP ^43^. Expression by transient transfection of CHO^BRI/55E1™^ cells was performed using a previously described high cell density method ^15^. Since a significant proportion of N was released from transfected cells despite high viability, it was purified from the culture supernatant. Following centrifugation and filtration, supernatant harvested 7 d post-transfection was purified by immobilized metal affinity chromatography (IMAC) on a Ni Sepharose Excel column (Cytiva, Vancouver, BC, Canada). The column was washed with 50 mM sodium phosphate buffer (pH 7.0) containing 25 mM imidazole and 300 mM NaCl, and N was eluted with 50 mM sodium phosphate buffer (pH 7.5) containing 300 mM imidazole and 300 mM NaCl. N was further purified by affinity chromatography on a StrepTrap XT Chromatography Column (Cytiva) equilibrated in Buffer W (100 mM Tris pH 8.0, 150 mM NaCl). The column was washed with Buffer W and bound protein was eluted with Elution Buffer (Buffer W supplemented with 50 mM biotin and 1 mM ethylenediaminetetraacetic acid (EDTA)). Purified N was buffer exchanged into Dulbecco’s PBS (PBS) using a CentriPure P100 Gel Filtration Column (Apex Scientific, Maynooth, Ireland), sterile-filtered through a 0.2 μm membrane, and stored at -80°C. For preparation of reference material aliquots (NCAP-1) for distribution by National Research Council of Canada (NRC) Metrology, the bulk purified protein was processed through an additional buffer exchange in 50 mM Tris-HCl pH 8.0 supplemented with 150 mM sodium chloride. N integrity and purity was analyzed by SDS-PAGE and analytical SEC-HPLC, which was run in PBS supplemented with 200 mM arginine on a 5 × 150 mm Superdex 200 HR column (Cytiva).

#### RBD

Amino acids 331–521 of the SARS-CoV-2 spike protein (YP_009724390.1, Wuhan-Hu-1 strain) were cloned into the pTT5® vector using EcoRI and BamHI. The construct encodes an N-terminal human interleukin 10 signal peptide (MHSSALLCCLVLLTGVRA) followed by a Twin-Strep-tag II-(His)_6_-FLAG tag fused to the RBD N-terminus. The construct was expressed by transient gene expression in CHO^BRI/55E1™^ cells as described above ^15^. Clarified culture supernatant harvested 8 d post-transfection was purified by IMAC on Ni Sepharose Excel columns as above. The IMAC eluate was buffer-exchanged using NAP-25 columns (Cytiva) into PBS before a second affinity purification step using Strep-Tactin XT Superflow (IBA Lifesciences, Göttingen, Germany), following the manufacturer’s instructions. The pooled eluate (de-salted into PBS and concentrated) was applied to a Superdex-75 gel filtration column (Cytiva). SEC fractions containing RBD with low levels of high-molecular-weight contaminants were pooled. The second RBD construct (RBD^319–541^) consists of amino acids 319–541 with a C-terminal (His)_6_-FLAG tag. The protein was expressed and purified as described for RBD^331–521^, except the Strep-Tactin XT purification step was omitted.

RBD and N antigens were compared against previously validated antigens produced by the Rini and Sicheri laboratories, respectively, and described in ^8, 13^.

#### Recombinant antibody production

VHH and mAb sequences were synthesized by GenScript using C. griseus codon bias and cloned into the pTT5® plasmid. The llama single domain antibody (VHH) VHH72 was described previously (PDB entry 6WAQ_1) ^17^. Additional VHHs (NRCoV2-04 and NRCoV2-20) were isolated in-house from llamas immunized with recombinant SARS-CoV-2 trimeric spike ectodomain SmT1 (Supplementary figure 2). VHH sequences were fused to an antibody-dependent cell-mediated cytotoxicity (ADCC)-attenuated human IgG1 Fc domain (hFc1X7, from patent US 2019 352 383A1) to generate VHH72-Fc, NRCoV2-04-Fc and NRCoV2-20-Fc. The anti-human-IgG monoclonal antibodies (mAbs) IgG#5 and IgG#6 were derived from mice immunized with human IgG; heavy chain (HC) and light chain (LC) variable domain sequences (VH and VL) were fused to mouse IgG2a and mouse kappa LC constant domain sequences, respectively, to express full-length mAbs. The HC was fused in-frame at its C-terminus with ferric HRP (PDB: 1W4W_A) to create IgG#5-HRP and IgG#6-HRP. These mAbs were also tested as HRP conjugates (rather than HRP fusions) as described in Supplementary figure 6. For protein production, VHH or mAb HC/LC (50:50 w/w) plasmids were transfected into CHO^BRI/55E1™^ cells using PEI-Max transfection reagent (Polysciences, Warrington, PA, USA) as described previously ^13, 15^. Clarified 0.2 μm-filtered supernatants were loaded on 5 mL HiTrap MabSelect SuRe columns (Cytiva) equilibrated in PBS, and the columns were washed with PBS prior to antibody elution with 100 mM citrate buffer, pH 3.6. The eluted antibodies were formulated in PBS by buffer exchange using NAP-25 columns. Purified proteins were quantified based on their absorbance at 280 nm and analyzed by analytical size-exclusion on an Acquity BEH200 column (Waters) by UPLC-MALS system (as described above for the spike trimer) or on a 5 × 150 mm Superdex 200 HR column (Cytiva) coupled to a high-performance liquid chromatography (SEC-HPLC) system (Waters).

#### Biotinylated ACE2 production

The human ACE2 (UniProtKB-Q9BYF1) cDNA was synthesized by GenScript and optimized for expression in CHO cells. The construct encodes a human interleukin 10 signal peptide (MHSSALLCCLVLLTGVRA) followed by a Twin-Strep-tag II-(His)_6_-FLAG tag on the N-terminus of the mature ACE2 receptor ectodomain (amino acids 20–613). A biotin acceptor peptide (BAP) sequence (GLNDIFEAQKIEWHE) was added in-frame at the ACE2 C-terminus. The cDNA was cloned into pTT5® using EcoRI and BamHI. The ACE2-BAP cDNA was expressed by transient gene expression in CHO^BRI/55E1™^ cells as described ^15^ with the addition of 5% (w/w) pTT5®-BirA (an *Escherichia coli* biotin ligase) expression plasmid as described previously ^44^. Clarified culture supernatant harvested 8 d post-transfection was purified by IMAC on Ni Sepharose Excel columns as described for N above. The IMAC eluate was buffer-exchanged using NAP-25 columns into PBS before a second affinity purification step using Strep-Tactin XT Superflow. The pooled eluate (buffer-exchanged into PBS as above) was stored at -80°C. Biotinylated ACE2 from the NRC was compared against biotinylated ACE2 produced in the Rini lab and previously described in^13^.

### Participant recruitment and study approval

All research was performed in accordance with Canada’s Tri-Council Policy Statement: Ethical Conduct for Research Involving Humans. External samples and data were transferred via Materia and Datal Transfer Agreements as appropriate. Samples were de-identified prior to transfer to the assay laboratory.

#### For Toronto cohorts

Negative control serum samples were from patients enrolled in cancer studies pre-COVID-19 (prior to November 2019; Mount Sinai Hospital (MSH) Research Ethics Board (REB) studies #01-0138-U and #01-0347-U), which were archived and frozen in the Lunenfeld-Tanenbaum Research Institute (LTRI) Biobank. Convalescent serum samples were obtained from in- and out-patients with polymerase chain reaction (PCR)-confirmed COVID-19 by the Toronto Invasive Bacterial Diseases Network in Toronto and the regional municipality of Peel, Ontario (REB studies #20-044 Unity Health Network, #02-0118-U/05-0016-C, MSH). Specimen-only Canadian Blood Services (CBS) serum donations were collected from individuals who met one or more of the following criteria: (a) indicated they had a SARS-CoV-2-positive PCR test, (b) a declaration of having been a close contact of a COVID-19 case, (c) a travel history and clinical presentation compatible with COVID-19, and (d) signs and symptoms compatible with COVID-19. ELISAs were conducted at the LTRI with MSH Research Ethics Board (REB) approval (study number: 20-0078-E). Samples from vaccinated individuals were obtained from Sunnybrook Health Sciences Centre (REB study #4814, Sunnybrook, #21-0049-E, MSH) and from MSH (REB study #20-0144-A).

#### For Ottawa cohorts

Pre-pandemic serum and plasma samples were collected between April 2015 and December 2019 from diverse sources, including the Eastern Ontario Regional Laboratory Association (EORLA) and the Ottawa Hospital (TOH). Pediatric samples were acquired from the BC Children’s Hospital Biobank (BCCHB) in Vancouver, BC, Canada (REB#: H-07-20-6009). Pandemic samples were collected from longitudinal studies of severe and mild hospitalized cases of COVID-19 (REB # H-04-20-5727 and # H-11-20-6172), and from a surveillance study of at-risk and convalescent individuals called *Stop the Spread Ottawa* (REB # H-09-20-6135) at the Ottawa Hospital Research Institute and the University of Ottawa.

### Sample collection, handling, and viral inactivation

#### Serum and plasma collection

Samples were collected using standard phlebotomy procedures at collection sites or self-collected by individuals after capillary puncture. In Toronto, inactivation of potential infectious viruses in plasma or serum was performed by incubation with Triton X-100 to a final concentration of 1% for 1 h prior to use ^8, 45^. Serum and plasma samples collected and processed in Ottawa did not require viral inactivation, as they were handled and tested within the University of Ottawa CL2+ biocontainment facility.

#### DBS preparation

Samples were prepared at the National Microbiology Laboratory of Canada (NML) as in ^46^. For panels 1–3, plasma from SARS-CoV-2 antibody-positive COVID-19 convalescent donors (MSH, Toronto) and SARS-CoV-2-negative donors (NML, Winnipeg) were used to generate matched plasma and contrived DBS samples. For contrived DBS samples, fresh SARS-CoV-2 antibody-negative blood was centrifuged, the plasma was removed, and the red blood cells were resuspended 1:1 in antibody-positive plasma and spotted (5 × 75 µL) onto Whatman 903 Protein Saver Cards (GE Healthcare, Boston, MA, USA), which were dried at room temperature overnight, then stored with desiccant in gas-impermeable bags at -80°C until testing. SARS-CoV-2 antibody-negative blood was spotted directly from EDTA Vacutainer tubes onto DBS cards. For panel 4, both positive and negative blood samples were spotted directly from EDTA tubes. All matched plasma and contrived DBS samples were tested using the anti-SARS-CoV-2 ELISA IgG kit (EUROIMMUN, Lübeck, Germany), according to the manufacturer’s instructions, to verify that donors were either positive or negative for SARS-CoV-2 antibodies prior to shipping to Toronto and Ottawa.

#### DBS handling: Toronto

Samples from DBS cards were punched manually using a 6 mm punch or in a semi-automated manner using a BSD600 Ascent puncher (BSD Robotics, Brisbane, QLD, Australia) with the indicated punch size. We first compared 6 mm circle punches from contrived DBSs and matching plasma samples (Supplementary figure 10) by eluting the punches in 250 μL PBST plus 1% Triton X-100 (0.226 mm^2^ µL^-1^ eluate). After this initial test, to ensure sufficient eluate to test all three antigens from 1 or 2 3-mm punches, we selected a final elution ratio of 0.176 mm^2^ µL^-1^ elution buffer (i.e. 80 μL for 2 × 3 mm punches). Eluted samples were centrifuged at 1,000 × g for 30 s before transfer to 96-well plates. Eluates were diluted 1:4 in 1.3% Blocker BLOTTO (Thermo Fisher Scientific, Waltham, MA, USA) to a final concentration of 1% in PBST, unless otherwise specified.

#### DBS handling: Ottawa

Samples from DBS cards were punched manually or in a semi-automated manner using a DBS puncher (PerkinElmer, Woodbridge, ON, Canada; 3.2 mm discs) or a BSD600 Ascent puncher (BSD Robotics; 3 mm discs) and eluted in 100 μL per disc PBS + 1% Triton X-100 for up to 16 h (minimum 4 h) in 96-well U-bottom plates on a shaker at room temperature. Elution optimization data are presented in Supplementary Figure 7. Eluates were centrifuged at 216 × g for 2 min and diluted 1:2 in 2% milk + PBST for a final concentration of 1% milk in PBST.

### Colorimetric direct ELISAs: Toronto protocol

Manual colorimetric ELISAs were adapted from assays described in ^8, 13^. Antigens (spike trimer, RBD 331–521, and N) at the indicated amounts (in ng, see Supplementary table 2 for optimized amounts) were diluted in 50 μL PBS and adsorbed onto a 96-well high-binding polystyrene Greiner Bio-One plate (Thermo Fisher Scientific, #655061) overnight at 4°C. Wells were washed three times with 200 µL PBST and then blocked with 200 µL 3% w/v milk powder (BioShop Canada Inc., Burlington, ON, Canada, #ALB005.250) in PBST for 1–2 h at room temperature. Samples were diluted as indicated in 50 µL of 1% w/v (final) milk powder in PBST and added to each plate. A standard curve of recombinant antibody was added to each plate in 50 µL 1% w/v milk powder in PBST at the indicated concentrations. For spike and its RBD, the recombinant antibodies used were VHH72-Fc (NRC; see above), human anti-spike S1 IgG (clone HC2001, GenScript, #A02038), human anti-Spike S1 IgM (clone hIgM2001, GenScript, #A02046), and human anti-spike IgA (clone CR3022, Absolute Antibody, Oxford, United Kingdom, #Ab01680-16.0). For N, the antibodies used were human anti-nucleocapsid IgG (clone HC2003, GenScript, #A02039), anti-nucleoprotein IgM (CR3018 (03-018), Absolute Antibody, #Ab01690 -15.0), and anti-nucleoprotein IgA (CR3018 (03-018), Absolute Antibody, #Ab01690 -16.0). Negative control antibodies purified from human serum (final 1 µg mL^-1^; human IgG, Sigma-Aldrich, Oakville, ON, Canada, #I4506; human IgM, Sigma-Aldrich #I8260, human IgA, Sigma-Aldrich, #I4036) and pools of positive and negative sera from 3–4 patients were added to each plate in 50 µL 1% w/v milk powder in PBST for quality control and to enable cross-plate comparisons. Samples were incubated for 2 h at room temperature, and wells were washed three times with 200 µL PBST. Anti-human secondary antibodies (recombinant anti-human IgG#5-HRP, goat anti-human IgG Fcy-HRP (Jackson ImmunoResearch Labs, West Grove, PA, USA, #109-035-098), goat anti-human IgM fc5u-HRP (Jackson ImmunoResearch Labs, #109-035-129), and goat anti-human IgA α chain-HRP (Jackson ImmunoResearch Labs, #109-035-127) were added to the plate at the indicated dilutions in 50 µL 1% w/v milk powder in PBST and incubated for 1 h at room temperature. Wells were washed three times with 200 µL PBST, then 50 μL of 1-Step Ultra TMB-ELISA Substrate Solution (Thermo Fisher Scientific, #34029) was added for 15 min at room temperature. The reaction was quenched with 50 μL Stop Solution for TMB Substrates (Thermo Fisher Scientific, #N600). The plates were read at 450 nm in a Cytation 3 Cell Imaging Multi-Mode Reader (BioTek Instruments Inc., Winooski, VT, USA). Blank values (the mean of the blanks in each 96-well plate) were subtracted from all raw reads prior to data analysis.

### Colorimetric direct ELISA: Ottawa protocol

Manual colorimetric ELISAs were modified from the assay described in ^47^. High protein-binding Immulon 4 HBX clear 96-well plates (VWR International, Mississauga, ON, Canada, #62402-959, note these plates have also been tested and validated in Toronto) were coated with 50 μL of 2 μg mL^-1^ antigen (spike, RBD 319–541, and N diluted in sterile PBS (WISENT Inc, St-Bruno, QC, Canada, #311-010-CL)) were incubated at 4°C on a shaker overnight. The next day, coated plates were washed three times with 200 μL of PBST and blocked with 200 μL of 3% w/v skim milk powder in PBST for 1 h at room temperature on a shaker at 700 rpm. The blocking buffer was removed, and plates were washed three times with 200 μL PBST. Serum and plasma samples were diluted in 1% w/v skim milk powder in PBST. An isotype-specific standard curve was included on each plate to enable cross-plate comparison: anti-SARS-CoV-2 S CR3022 Human IgG1 (Absolute Antibody, Ab01680-10.0), anti-SARS-CoV-2 S CR3022 Human IgA (Absolute Antibody, Ab01680-16.0), or anti-SARS-CoV-2 S CR3022 Human IgM (Absolute Antibody, Ab01680-15.0). Serum/plasma samples, standard curve, positive and negative controls, and blanks (100 μL/well in 1% w/v skim milk in PBST) were added and incubated for 2 h at room temperature on a plate shaker at 700 rpm. The plates were washed four times with 200 μL PBST, HRP-linked secondary antibody diluted in 1% milk in PBST (50 μL) was added, and the plate was incubated for 1 h at room temperature on a shaker at 700 rpm. The final isotype-specific secondary antibodies used were anti-human IgG#5-HRP (Supplementary figure 6), anti-human IgA-HRP (Jackson ImmunoResearch Labs, 109-035-011), and anti-human IgM-HRP (Jackson ImmunoResearch Labs, 109-035-129). Plates were washed four times with 200 μL PBST and developed using 100 μL of SIGMAFAST OPD Tablets (Sigma-Aldrich, P9187) dissolved in 20 mL Gibco Water for Injection for Cell Culture (Thermo Fisher Scientific, A1287301, final concentration of 0.4 mg mL^-1^ OPD, 0.4 mg mL^-1^ urea hydrogen peroxide, and 0.05 M phosphate-citrate, pH 5.0). After 10 min of incubation in the dark, the reaction was stopped with 50 μL of 3 M HCl and the absorbance was measured at 490 nm using a PowerWave XS2 Plate Reader (BioTek Instruments). Wells filled with dilution buffer were used as background controls and their reads were subtracted from serum values. Colorimetric assay optimization data are presented in Supplementary figures 5 and 6.

### Chemiluminescent direct ELISA: Toronto protocol and optimization

Automated chemiluminescent ELISAs were adapted from assays first described in ^8^, and performed using liquid dispensers (Biomek NXp (Beckman, Indianapolis, IN, USA), Multidrop Combi (Thermo Fisher Scientific)) and a washer (405 TS/LS LHC2 (Biotek Instruments)); all wash steps included four washes with 100 μL PBST) on a Thermo Fisher Scientific F7 Robot System at the Network Biology Collaborative Centre (nbcc.lunenfeld.ca). All incubations were performed at room temperature. Antigen (spike trimer, RBD 331–521, or N) at the indicated amounts (ng) were diluted in 10 μL PBS and dispensed into the wells of a 384-well LUMITRAC 600 high-binding polystyrene Greiner plate (Thermo Fisher Scientific, #781074). The plate was centrifuged at 233 × g for 1 min to ensure even coating, incubated overnight at 4°C, and washed. Wells were blocked with 80 µL of 5% Blocker BLOTTO for 1 h and then washed. Samples and controls (as in the colorimetric assay) were diluted as indicated in a final concentration of 1% Blocker BLOTTO in PBST, and 10 µL was added to each well from 96-or 384-well source plates. Plates were incubated for 2 h, and wells were washed with PBST. Secondary antibodies (as in the colorimetric assay) were diluted as indicated in 1% Blocker BLOTTO in PBST and 10 µL was added to each well. After incubation for 1–2 h, the wells were washed and 10 µL of ELISA Pico Chemiluminescent Substrate (Thermo Fisher Scientific, #37069, diluted 1:4 in ddH_2_0) was dispensed into each well and mixed at 900 rpm for 10 s. After a 5–20-minute incubation, plates were read on an EnVision 2105 Multimode Plate Reader (PerkinElmer) at 100 ms/well using an ultra-sensitive luminescence detector.

Protein reagents from the NRC (described above) were first optimized in the 96-well colorimetric manual assay using recombinant antibodies and serum samples from CBS (Supplementary figure 16). The final amount of antigen per well was then scaled down by 3.5–4-fold to migrate to 384-well format for the automated assay. Two concentrations of secondary antibody IgG#5-HRP (0.09 and 0.18 μg mL^-1^) were assessed using dilution curves of the VHH72-Fc antibody (to detect spike and its RBD) or an anti-N antibody (to detect N; Supplementary figure 17), and the best concentration (0.18 μg mL^-1^) was further tested on a dilution series of 32 serum samples provided by CBS (Supplementary figure 3). We also tested anti-RBD NRCoV2-04-Fc and NRCoV2-20-Fc recombinant calibration antibodies, which were comparable to VHH72-Fc as reference curves (Supplementary figure 17). For IgA and IgM detection, reagents were first tested in the colorimetric assay (Supplementary figure 8). For chemiluminescent assays, 10 µL of goat anti-human IgM-HRP (1:10,000; 0.80 ng/well) or goat anti-human IgA-HRP (1:12,000, 0.66 ng/well) were used as secondary antibodies.

### Chemiluminescent direct ELISAs: Ottawa protocol and optimization

Automated chemiluminescent ELISAs were based upon and optimized from assays first described in^48^, and performed using MicroLab Star robotic liquid handlers (Hamilton, Reno, NV, USA) and a 405 TS/LS LHC2 plate washer (Biotek Instruments; all wash steps included four washes with 100 μL PBST) at the University of Ottawa, Faculty of Medicine (Roger Guindon Hall). All incubations were done at room temperature with shaking at 500–700 rpm. Antigens (spike, RBD 319–541, and N) were diluted in PBS and dispensed into the wells of a 384-well high-binding polystyrene Nunc plate (Thermo Fisher Scientific, #460372) at a final amount of 50 ng/well. The plates were centrifuged at 216 × *g* for 1 min to ensure even coating, incubated overnight rocking at 4°C, and washed. Wells were blocked with 80 µL of 3% w/v skim milk powder dissolved in PBST for 1 h and then washed. Samples and controls were diluted as indicated to a final concentration of 1% w/v skim milk powder in PBST and 10 µL was added to each well from a 96-well source plate. Plates were incubated for 2 h and wells were washed. Secondary antibodies (as in the colorimetric assay) were diluted as indicated in 1% w/v skim milk powder in PBST and 10 µL was added to each well. After incubation for 1 h, the wells were washed and 10 µL of ELISA Pico Chemiluminescent Substrate (diluted 1:2 in MilliQ H_2_O) was dispensed into each well. After a 5 min incubation with shaking, plates were read on an Neo2 plate reader (BioTek Instruments) at 20 ms/well and a read height of 1.0 mm.

### Colorimetric snELISAs: Toronto protocol and optimization

The snELISA assay was performed as described ^13^ with the indicated antigens and ACE2. All wash steps included four washes in 200 μL PBST. RBD 319–541 or spike was adsorbed onto 96-well high-binding polystyrene Greiner Bio-One plates (Thermo Fisher Scientific, #655061) at 100 or 200 ng/well, respectively, in 50 µL PBS and incubated overnight at 4°C. Plates were washed, then blocked for 1–1.5 h at room temperature with 200 μL 3% bovine serum albumin (BSA; BioShop Canada Inc., SKI400.1). After washing, serum or plasma was added to the plate at the indicated concentrations in 50 µL 1% BSA in PBST (final concentration) and incubated for 1 h. Wells were washed and incubated with 50 µL of biotinylated recombinant ACE2 as indicated for 1 h. After washing, wells were incubated with 44 ng Streptavidin-Peroxidase Polymer, Ultrasensitive (Sigma-Aldrich, S2438) for 1 h. The resultant signal was developed and quantified with TMB-ELISA in an identical manner to the colorimetric direct ELISAs. Due to day-to-day variations in signal, all optical density at 450 nm (OD_450_) values were normalized to the OD_450_ of the well without serum or antibody for each sample. All values are expressed as ratios.

### Chemiluminescent snELISAs: Toronto protocol

The automated snELISA assay was performed on the same F7 platform as the direct detection ELISA with the same washing protocol and incubation temperature. Greiner 384-well Lumitrac 600 plates were coated with RBD 319–541 (34 ng/well) or spike (50 ng/well) following the same protocol as for direct detection. Before each of the next four steps, the plates were washed four times with PBST: 1) the wells were blocked with 80 μL 3% BSA in PBST for 1 h; 2) Plasma or serum sample was dispensed at the indicated dilutions in 10 or 20 μL and incubated for 2 h; 3) 10 μL of ACE2-BAP (2.08 ng/well in 1% BSA in PBST) or ACE2-Rini (17 ng/well in 1% BSA in PBST) was added to each well and the plates were incubated for 1 h; 4) 10 μL of Streptavidin-Peroxidase Polymer, Ultrasensitive (15 ng/well in 1% BSA in PBST) was added to each well and the plates were incubated for 1 h. Addition of ELISA Pico Chemiluminescent Substrate and reading on the EnVision 2105 Multimode Plate Reader (PerkinElmer) were performed as for direct detection. All values were normalized to blanks (with no samples added) on the same 384 well plate.

### Chemiluminescent snELISAs: Ottawa protocol

The surrogate neutralization ELISA first described in ^13^ was adapted and optimized for compatibility with the Hamilton MicroLab STAR robotic liquid handler. The methods for plate washing steps, incubations, and data acquisition were as described for the automated chemiluminescent ELISA. 384-well high-binding polystyrene Nunc plates (Thermo Fisher Scientific, #460372) were coated with 100 ng/well of spike or RBD 319–541, centrifuged in a plate spinner to ensure even coating, and incubated overnight with rocking at 4°C. Plates were washed, 80 μL of blocking solution (3% w/v skim milk powder in PBST) was added to each well, and the plates were incubated for 1 h at room temperature on a shaker at 700 rpm. During the blocking step, serum/plasma samples or DBS eluates were diluted in skim milk powder in PBST to a final milk concentration of 1% w/v. For single-point neutralizations, samples were diluted 1:5, or to calculate the half maximal effective concentration (EC50), the samples were titrated using a 5-point curve (1:5, 1:25, 1:125, 1:625, 1:6,250). To facilitate quality control, downstream analysis, and cross-plate comparisons, standard curves of NRCoV2-20-Fc and three dilutions (1:5, 1:125, and 1:6,250) of pooled positive and negative serum samples were included in quadruplicate on each plate. Serum-free/ACE2-free and serum-free wells were also included on each plate to establish the minimum and maximum signals, respectively. Plates were washed, and samples and controls (20 µL) were added to the wells and incubated for 2 h. Plates were then washed, and 6.5 ng of ACE2-BAP diluted in 1% w/v skim milk powder in PBST was added to each well and incubated for 1 h. Plates were again washed, and Streptavidin-Peroxidase Polymer (Sigma #S2438), diluted in 1% w/v skim milk powder in PBST at a concentration of 1.25 ng μL^-1^ (25 ng/well) was added to each well and incubated for 1 h. Plates were washed a final time and 20 µL of ELISA Pico Chemiluminescent Substrate (diluted 1:2 in MilliQ H_2_0) was dispensed into each well. After a 5-min incubation, plates were read on an Neo2 plate reader (BioTek Instruments) at 20 ms/well and a read height of 1.0 mm. Assay optimization data are presented in Supplementary Figure 14.

### Data analysis: Toronto

#### Quality control and determination of relative ratios

Raw values for each sample (luminescence counts per second) were normalized to a blank-subtracted reference point from the reference curve (0.0156 μg mL^-1^ for VHH72-Fc (for spike/RBD) or 0.0625 μg mL^-1^ for anti-N (for N)) to create relative ratios. For each automated test, the raw values and relative ratios of each control were compared to those of prior tests to confirm their similarity. The distributions of the raw values of the reference points and positive controls compared to the blanks and the reference points for each antigen compared to each other should be within the 90% confidence ellipse of the distributions of previous tests. In addition, the reference points should be within 15% coefficient of variation (CV) of prior runs. The log_10_ sample density distributions of the raw values and relative ratios were compared to prior runs to confirm that they were within range. For points outside of these ranges, the individual control was either removed (e.g. if it was a single outlier out of the four on the plate) or the plate or test was repeated.

#### Receiver-operating characteristic (ROC) analysis

Samples acquired prior to November 2019 (pre-COVID) were considered true negatives and samples from convalescent patients with PCR-confirmed COVID-19 were considered true positives (for IgM and IgA analyses, we selected positives that were collected 20–40 d post-symptom onset). ROC analysis using relative ratios was performed with the pROC package in R ^49^ with default parameters. For IgG (serum/plasma and DBS), the analysis was performed in duplicate, with individual replicates considered individual samples. To avoid having samples with technical errors skew the threshold determination, negative plasma samples that were positive in only one replicate and negative DBS samples whose values were more than four standard deviations (SDs) from the mean of the log distribution of negative samples were removed from the analysis.

#### Thresholding based on the control mean

For plasma and serum IgG, negative controls (blanks, IgG pools, 0.25 and 0.0625 μL/well of a negative master mix (including equal volumes of four pre-COVID-19 samples (N2, N11, N39, N41), n = 1,320 for spike, 1,248 each for RBD and N) from 23 experiments over 4 months were included in the analysis. We used three SDs from the mean of the log_10_ distribution of the relative ratios of these negative controls as the threshold for each antigen. The same strategy was employed for IgA (72 negative controls from two experiments 1 month apart) and IgM (80 negative controls from three experiments over 2 months).

#### Scoring positives

Positive and negative results were first defined for each antigen, based on the defined thresholds. For projects where an overall assessment of the confidence of antibody detection in a sample is required, we imposed a sample-level rule: the sample must have passed positivity in at least 2 out of 3 antigen tests.

#### Other data analyses

Plots were generated in R using the ggplot2, lattice, latticeExtra, grid, and gridExtra packages. Correlations between samples were calculated using the cor() function in the R stats package, using the Spearman method. The density distributions of the log of the signals scaled to the reference were separated using the normalmixEM mixtool function in the R mixtools package, based on an expected minimization of only two subpopulations assumed to have a Gaussian distribution. normalmixEM initialization, mean, and SD parameters were based on visual estimation of the density distribution of the log of the signal scaled to the reference. The initially assumed proportion between the two subpopulations was set to 50%. Density distributions were plotted using default R parameters (bandwidth: bw.nrd0, which implements a rule-of-thumb for choosing the bandwidth of a Gaussian kernel density estimator).

#### Calibration to the WHO standard for antibody detection

For each antigen, the relative ratio from the WHO International Standard (IS, National Institute for Biological Standards and Control (NIBSC, South Mimms, United Kingdom), Code 20/136, pooled convalescent plasma) at different sample dilutions (in binding antibody units (BAU) mL^-1^) was represented in log-log scale. The response curve was modelled by an S-shaped sigmoid curve (y = a*x/(1+b*x)+c, expressed in log(y), log(x) coordinates). The nls function (nonlinear least squares) from the R stats package was used to best match the response to the measured data. The interval where the log response was considered linear to the log of the BAU mL^-1^ was selected visually. The lm (linear model) function from the R stats package was used to obtain the parameters of the linear approximation of the curve (Figure 3a). The WHO International Reference panel (NIBSC code 20/268), which contains pooled plasma samples that are Negative (code 20/142), Low (code 20/140), Low S High N (code 20/144), Mid (code 20/148), and High (code 20/150), was measured at different concentrations. Estimated BAU mL^-1^ values were obtained from the above linear approximation using only sample dilutions that were within this linear range, and further adjusted by accounting for the dilution factor. To convert from relative ratios (RRs) to BAU mL^-1^ for plasma or serum samples where only the reference curve was included in the same test, (Figure 3d), a conversion formula can be applied:

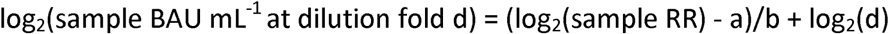

where a and b represent the y-intercept and slope of the linear interval of the IS curve (Supplementary table 4).

#### Calibration to the WHO standard for snELISA

The WHO IS and the WHO International Reference Panel were analyzed by snELISA at the indicated dilutions to convert values to International Units (IU mL^-1^). The same approach was applied as for the antibody detection ELISA except we used the sample relative ratio (RR) instead of the log_2_ of the sample RR because the linear range was larger. We then applied this formula:

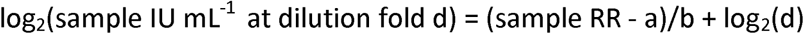

where a and b represent the y-intercept and slope of the linear interval of the IS curve respectively and d is the dilution factor of the sample. For RBD as antigen, a = 1.58 and b = -0.16.

### Data analysis: Ottawa

For consistency in data processing, a constant plate layout was established, containing all controls and standard curves in quadruplicate (Supplementary figure 11a). Luminescence values obtained from the isotype- and antigen-specific standard curve were modeled using a four-parameter log-logistic function to identify the inflection point (Supplementary figure 11b). Blank-subtracted luminescence values were scaled in relation to the curve to allow data normalization for subsequent processing. Using a 3% false discovery rate (FDR) calculated using a density distribution from a screen of adult and pediatric pre-pandemic samples, a threshold was established for each isotype-antigen combination. Signal to threshold (also known as signal-to-cutoff, S/CO) ratios specific to each isotype-antigen combination were then calculated and sample positivity was determined (Supplementary figure 11c–d). For example, for IgG, both spike and N antigens were considered. If respective antigens were detected over the established threshold (S/CO ≥ 1), a positive call was made. All analyses were performed in R. Density distributions were determined using the default density function with default parameters. Standard curve processing was performed using the LL.4 four-parameter log-logistic self-starter function from the drc package ^50^, also with default parameters. Plots were generated using the ggplot2 and reshape2 packages.

Using NML panel 4, containing known positive and negative DBS samples, the optimal FDR threshold value for SARS-CoV-2 IgG-positive calls was determined by testing FDRs of 1–100% and calculating the number of true positive (TP), false positive (FP), true negative (TN), and false negative (FN) samples. Maximum accuracy occurred at an FDR of 3% (Supplementary figure 11e). ROC curves and associated area under the curve measurements were generated in R using the pROC package with default parameters (Table 2, Supplementary figure 11f–h).

To convert luminescence values into BAU ml^-1^, the WHO IS was titrated for spike, RBD, and N (Supplementary figure 9). Scaled luminescence was calculated, as above, and response curves were modelled using the LL.4 four-parameter log-logistic self-starter function from the drc package with default parameters. The interval where the scaled luminescence response curves were considered linear to BAU was selected visually. Calibration of the three antigens was performed using the NIBSC 20/268 reference panel. The following formula was used to convert scaled luminescence values to BAU mL^-1^:

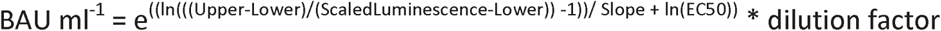

For snELISA, the WHO IS was titrated and ACE2-spike binding inhibition was reported as a function of IUs. Blank adjusted luminescence values were converted to inhibition of ACE2-spike binding; maximal signal (i.e. 0% inhibition) was measured in absence of convalescent plasma (1% w/v skim milk powder in PBST). The normalized data was fitted using the LL.4 four-parameter log-logistic self-starter function from the drc package with default parameters in R to correlate binding inhibition to IUs. The following formula was used to convert binding inhibition % to IU ml^-1^:

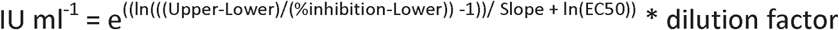

### Reagent access

The reagents produced by the NRC and qualified as reference materials (spike [SMT1-1], N [NCAP-1] and hACE2-BAP) are available through the Metrology Research Centre Virtual Store (<u>Virtual Store - National Research Council Canada: Products in Category Proteins (nrc-cnrc.gc.ca))</u>. A limited panel of matched plasma and DBS is available upon request

## Data Availability

This is a methods development manuscript.
REAGENT ACCESS: The reagents produced by the NRC and qualified as reference materials (SMT1-1, N and hACE2-BAP) are available through the Metrology Research Centre Virtual Store (Virtual Store - National Research Council Canada: Products in Category Proteins (nrc-cnrc.gc.ca)). A limited panel of matched plasma and DBS is available upon request

## Code availability

Code is available upon request.

## ACKNOWLEDGMENTS

We thank Alex Pelletier, Denis L’Abbé, Mélanie Duchesne, Simon Lord-Dufour, Manon Ouimet, Brian Cass, Louis Bisson, Alina Burlacu, Jean-Sébastien Maltais, Sergio Alpuche-Lazcano and other members of the NRC-HHT Mammalian Cell Expression Section for their contribution to the cloning, expression and purification of the various recombinant proteins used in this study. The contribution of Joline Cormier to recombinant protein sample management is greatly appreciated. We are grateful to Joe Schrag, Roger Maurice, and Mathieu Coutu from NRC-HHT (Montreal) for SEC-UPLC and SEC-HPLC analyses. We thank Jamshid Tanha and Martin Rossotti from NRC-HHT (Ottawa) for providing NRCoV2-04-hFc1X7 and NRCoV2-20-hFc1X7 antibodies. We thank Frank Sicheri and Derek Ceccarelli (LTRI) for contributing bacterially expressed N and James Rini (University of Toronto) for contributing RBD and biotinylated ACE2 protein to compare with the reagents described here. We also thank Lauren Caldwell for preprocessing some of the data generated from automated ELISAs. We thank Bruce Mazer, Mel Krajden and members of the COVID-19 Immunity Task Force (CITF) for coordinating immune response studies across the country and for supporting assay development and implementation. This is NRC’s publication #NRC-HHT_53561A. We acknowledge support from Ontario Together and funding from the Canadian Institutes of Health Research (CIHR; #VR1-172711, GA1-177703 and #439999) and the COVID-19 Immunity Task Force (CITF). We also acknowledge financial support from the NRC’s Pandemic Response Challenge Program. Funding for initial assay development in the Gingras lab was provided through generous donations from the Royal Bank of Canada and the Krembil Foundation to the Sinai Health System Foundation. The robotics equipment used is housed in the Network Biology Collaborative Centre at the LTRI, a facility supported by the Canada Foundation for Innovation, the Ontario Government, and Genome Canada and Ontario Genomics (OGI-139). Assay development in the Langlois lab and the purchase of robotic equipment was supported by a grant from the CIHR (#VR2-172722 and VS2 - 175569) and supplemental funding from the CITF. Kento T Abe was a recipient of an Ontario Graduate Scholarship and is supported by a CIHR CGS-D studentship. Yannick Galipeau is supported by a CIHR CGS-M studentship. Mariam Maltseva holds a Queen Elizabeth II Graduate Scholarship in Science and Technology (QEII-GSST). Anne-Claude Gingras is the Canada Research Chair in Functional Proteomics, and Marc-André Langlois, Anne-Claude Gingras, Jeffrey Wrana, and Samira Mubareka are members of CoVaRR-Net, the CIHR Coronavirus Variants Rapid Response Network.

## Competing interests

The authors declare no competing interests for this manuscript.

## Contributors

Karen Colwill, Anne-Claude Gingras, Marc-André Langlois, and Yves Durocher conceived the study. Karen Colwill, Anne-Claude Gingras, Marc-André Langlois, Matthew Stuible, Christian Gervais, Yannick Galipeau, Corey Arnold, and Yves Durocher wrote the paper. Yves Durocher, Matthew Stuible and Christian Gervais designed, performed, and validated reagent purification procedures. Linda Bennett, Kevin Yau, Angel X Li, Aimee Paterson, Michelle A Hladunewich, Pamela J Goodwin, Samira Mubareka, Allison J McGeer and Steven J Drews contributed essential patient samples. Bhavisha Rathod was responsible for sample intake at the Toronto site. François Cholette, Christine Mesa prepared and helped evaluate DBS panels and John Kim supervised the DBS panel evaluation. Yannick Galipeau, Corey Arnold, Bhavisha Rathod, Jenny H Wang, Mariam Maltseva, Mariam Iskilova, and Mahya Fazel-Zarandi performed serology experiments. Karen Colwill, Miriam Barrios-Rodiles, Kento T Abe, Martin Pelchat, Lynda Rocheleau, Yannick Galipeau and Adrian Pasculescu performed data analysis. Martin Pelchat and Adrian Pasculescu performed statistical modelling. Karen Colwill, Kento T Abe, Adrian Pasculescu, Yannick Galipeau, Mariam Maltseva, Lynda Rocheleau, Matthew Stuible and Martin Pelchat generated figures for the paper. Miriam Barrios-Rodiles and Jeffrey L Wrana provided oversight of the Toronto screening facility.

All the authors gave final approval of the version to be published and agreed to be accountable for all aspects of the work. Marc-André Langlois, Anne-Claude Gingras, and Yves Durocher are co-senior corresponding authors.

## Data Sharing

Raw and normalized values from the automated assays are provided in Supplementary table 5. R scripts used to analyze the data are available upon request.

## Supplementary figures

Supplementary figure 1 – Chromatogram traces for the purified ELISA antigens and detection reagents.

Supplementary figure 2 – Purification of recombinant anti-spike antibodies.

Supplementary figure 3 – Titration curves of 32 serum samples from CBS using automated ELISAs to detect (a) spike, (b) RBD 331–521, and (c) N antibodies.

Supplementary figure 4 – Performance of the IgG chemiluminescent ELISAs with plasma and serum.

Supplementary figure 5 – Comparison between o-Phenylenediamine dihydrochloride (OPD) and 3,3⍰,5,5⍰-tetramethylbenzidine (TMB) as colorimetric substrates for the manual 96-well plate ELISA.

Supplementary figure 6 – Comparison of IgG-HRP secondary antibody performance (a) Analysis of NRC IgG-HRP secondary (detection) antibodies.

Supplementary figure 7 – Elution and secondary antibody optimizations for IgG ELISAs using DBSs.

Supplementary figure 8 – IgA and IgM ELISAs.

Supplementary figure 9 – Conversion of serology data to WHO BAUs and validation of SARS-CoV-2 IgG profiling performed on the uOttawa automated serology platform.

Supplementary figure 10 – DBS optimization.

Supplementary figure 11 – Processing and validation of the Ottawa automated serology platform.

Supplementary figure 12 – ROC curve for panel 4 DBS IgG analysis.

Supplementary figure 13 – Optimization and testing of NRC reagents for snELISA.

Supplementary figure 14 – Optimization of the snELISA for the automated platform.

Supplementary figure 15 – Optimized snELISA for the automated platform.

Supplementary figure 16 – Optimization of the NRC reagents.

Supplementary figure 17 – Optimization of IgG#5-HRP and recombinant antibodies for chemiluminescent assays

## Supplementary Tables

Supplementary table 1 – Reagent information.

Supplementary table 2 – Comparison of manual and automated ELISAs.

Supplementary table 3 – ELISA performance statistics for plasma or serum IgA and IgM (Toronto).

Supplementary table 4 –Conversion of relative ratios to international units (BAU mL^-1^) for plasma or serum.

Supplementary table 5 – Raw data from automated experiments.

## Notes

### Competing Interest Statement

The authors have declared no competing interest.

### Author Declarations

The Research Ethics Board of Mount Sinai Hospital (Toronto, ON) gave ethics approval for performing ELISA assays with samples collected across several studies (study number: #20-0078-E). The Research Ethics Board of Mount Sinai Hospital also gave ethics approval for sample collection and analysis (studies #02-0118-U/05-0016-C, #01-0138-U, #20-0144-A, #21-0049-E and #01-0347-U). The Research Ethics Board of the Unity Health Network (Toronto, ON) gave ethics approval for sample collection and analysis (study #20-044). The Research Ethics Board of Sunnybrook Health Sciences Centre gave ethics approval for sample collection and analysis (study #4814). The British Columbia Childrens Hospital Biobank (BCCHB) in Vancouver, BC, Canada gave approval sample collection and analysis (REB#: H-07-20-6009). The University of Ottawa gave ethics approval for sample collection and analysis (REB # H-04-20-5727, # H-11-20-6172 and REB # H-09-20-6135).

### Summary of Updates

1. Changed the title and the focus of the manuscript to describe the assay to an international audience. 2. Expanded the assay conversion to WHO international units. This was also associated with extensive text editing.

