## Supplementary Figures for "A scalable serology solution for profiling humoral immune responses to SARS-CoV-2 infection and vaccination"

**Spike (SMT1)**

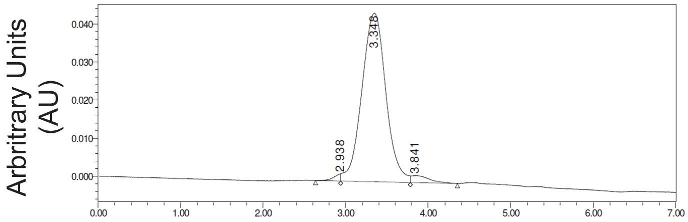


Minutes

Sample Conc. (mg mL$^{-1}$): 0.80

UPLC-SEC, BEH450

| | RT | % Area | Area | Height |
|---|---|---|---|---|
| 1 | 2.938 | 1.17 | 10935 | 1914 |
| 2 | 3.348 | 95.76 | 894783 | 44280 |
| 3 | 3.841 | 3.07 | 28719 | 1839 |

**RBD (331–521)**

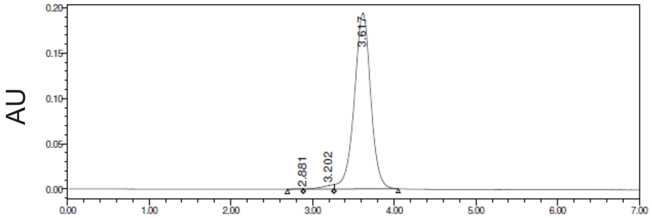


Minutes

Sample Conc. (mg mL$^{-1}$): 1.07

UPLC-SEC, BEH200

| | RT | % Area | Area | Height |
|---|---|---|---|---|
| 1 | 2.881 | 0.05 | 1376 | 269 |
| 2 | 3.202 | 1.63 | 44817 | 3980 |
| 3 | 3.617 | 98.32 | 2709774 | 193790 |

**RBD (319–541)**

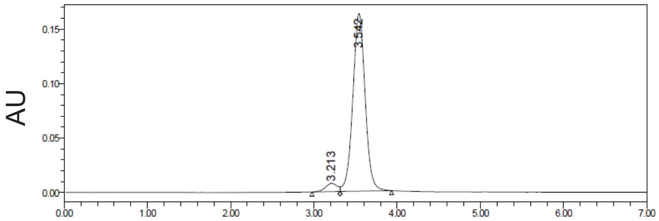


Minutes

Sample Conc. (mg mL$^{-1}$): 0.98

UPLC-SEC, BEH200

| | RT | % Area | Area | Height |
|---|---|---|---|---|
| 1 | 3.213 | 4.25 | 74411 | 7654 |
| 2 | 3.542 | 95.75 | 1675802 | 163023 |

**Nucleocapsid (NCAP)**

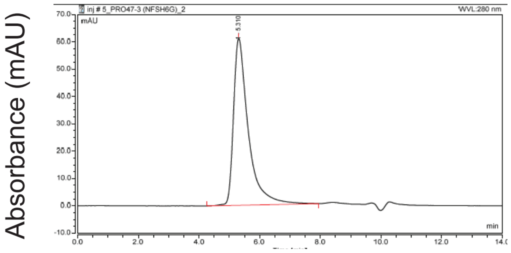


Minutes

Sample Conc. (mg mL$^{-1}$): 1.4

HPLC-SEC, Superdex

| | RT | % Area | Area | Height |
|---|---|---|---|---|
| 1 | 5.31 | 100 | 2070500 | 61109 |

**ACE2-BAP**

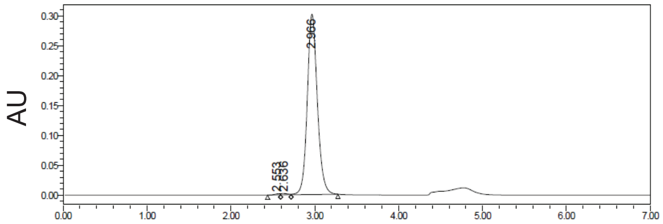


Minutes

Sample Conc. (mg mL$^{-1}$): 0.80

UPLC-SEC, BEH200

| | RT | % Area | Area | Height |
|---|---|---|---|---|
| 1 | 2.553 | 0.34 | 8581 | 1746 |
| 2 | 2.636 | 0.52 | 13284 | 2203 |
| 3 | 2.966 | 99.14 | 2521768 | 301927 |

**IgG#5-HRP**

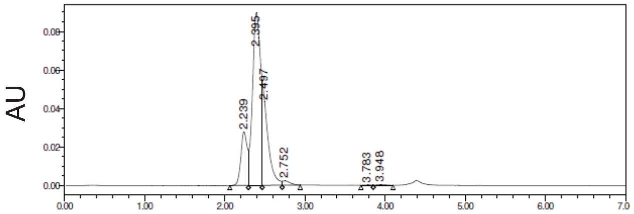


Minutes

Sample Conc. (mg mL$^{-1}$): 0.60

UPLC-SEC, BEH200

| | RT | % Area | Area | Height |
|---|---|---|---|---|
| 1 | 2.239 | 14.43 | 151481 | 27895 |
| 2 | 2.395 | 59.64 | 625983 | 89609 |
| 3 | 2.497 | 24.12 | 253149 | 43384 |
| 4 | 2.752 | 1.44 | 15156 | 2291 |
| 5 | 3.783 | 0.1 | 999 | 206 |
| 6 | 3.948 | 0.27 | 2835 | 369 |

**Supplementary figure 1.** Chromatogram traces for the purified ELISA antigens and detection reagents.

**(a)** **VHH72-Fc1X7**

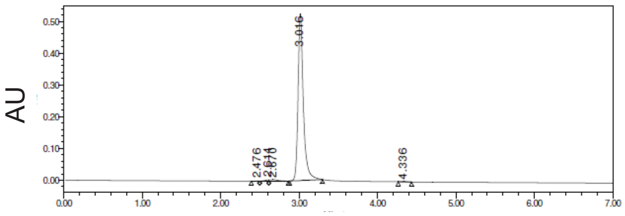


Minutes
Sample Conc. (mg mL$^{-1}$): 2.90

UPLC-SEC, BEH200

| | RT | % Area | Area | Height |
|---|---|---|---|---|
| 1 | 2.476 | 0.06 | 1401 | 407 |
| 2 | 2.614 | 0.48 | 12207 | 3640 |
| 3 | 2.670 | 1.27 | 31895 | 4615 |
| 4 | 3.016 | 97.93 | 2465652 | 524363 |
| 5 | 4.336 | 0.26 | 6552 | 1540 |

**(b)**

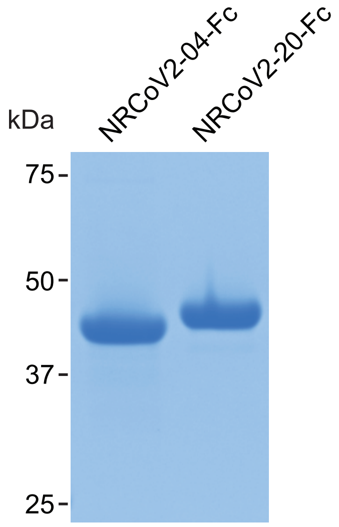


**(c)** **NRCoV2-04-Fc1X7**

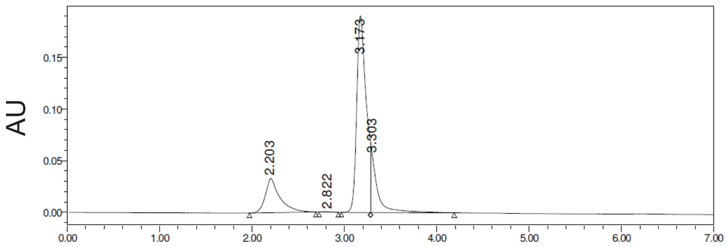


Minutes
Sample Conc. (mg mL$^{-1}$): 3.10

UPLC-SEC, BEH200

| | RT | % Area | Area | Height |
|---|---|---|---|---|
| 1 | 2.203 | 17.82 | 359486 | 33096 |
| 2 | 2.822 | 0.12 | 2507 | 422 |
| 3 | 3.173 | 66.68 | 1344847 | 190239 |
| 4 | 3.303 | 15.37 | 310017 | 55046 |

**NRCoV2-20-Fc1X7**

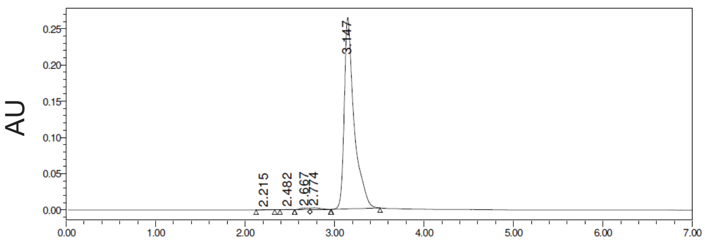


Minutes
Sample Conc. (mg mL$^{-1}$): 4.07

UPLC-SEC, BEH200

| | RT | % Area | Area | Height |
|---|---|---|---|---|
| 1 | 2.215 | 0.07 | 1525 | 259 |
| 2 | 2.482 | 0.05 | 1031 | 218 |
| 3 | 2.667 | 0.69 | 14239 | 2136 |
| 4 | 2.774 | 0.84 | 17367 | 2202 |
| 5 | 3.147 | 98.35 | 2030475 | 263884 |

**Supplementary figure 2.** Purification of recombinant anti-spike antibodies. **(a)** Chromatogram trace for VHH72-Fc. **(b)** Two additional recombinant anti-spike antibodies were analyzed on Coomassie-stained polyacrylamide gels under reducing conditions to assess their purity. Molecular weight markers (kDa) are shown to the left of the gel. **(c)** Chromatogram traces for the two additional anti-spike antibodies.

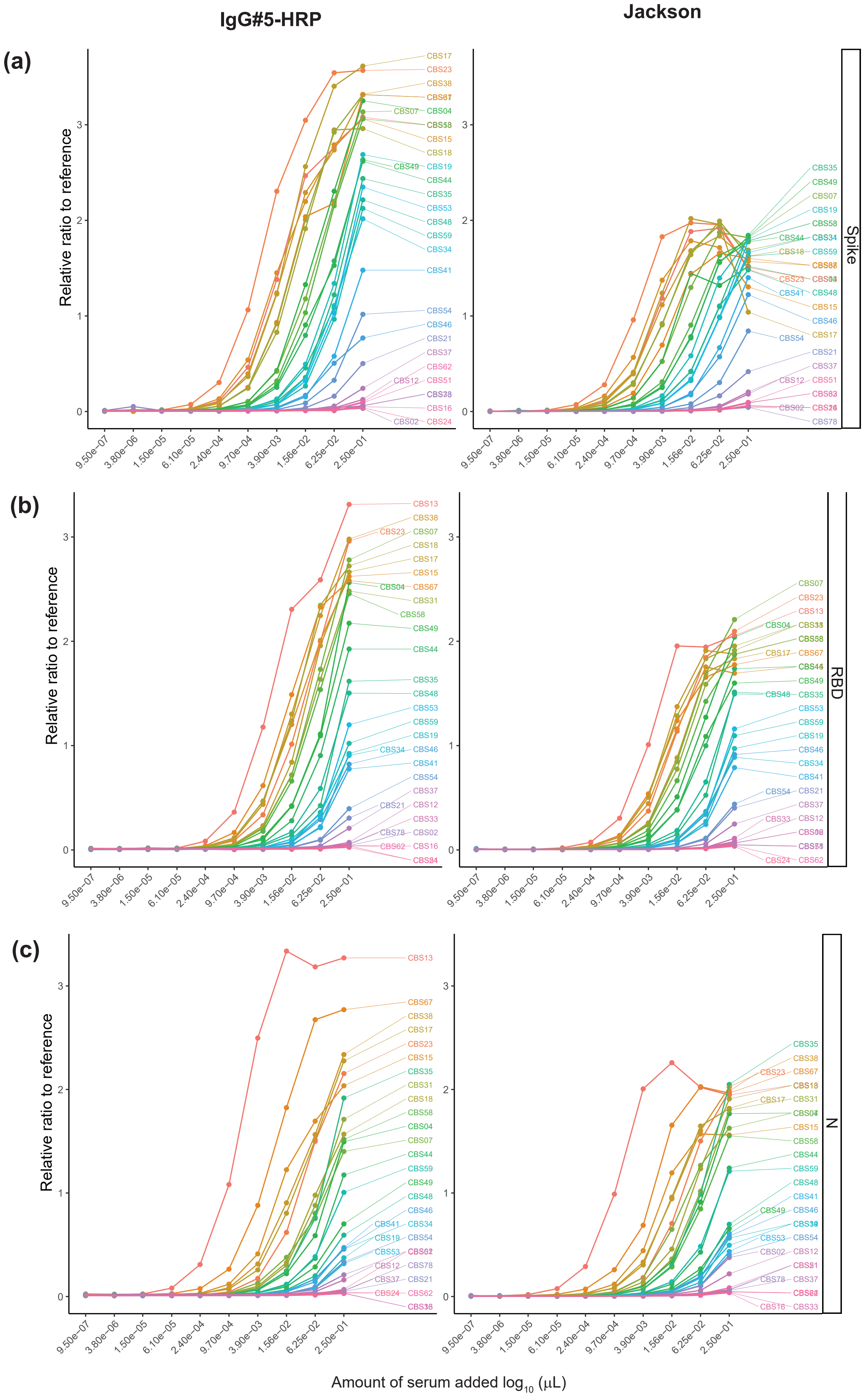


**Supplementary figure 3.** Titration curves of 32 serum samples from CBS using automated ELISAs to detect **(a)** spike, **(b)** RBD 331–521, and **(c)** N antibodies. IgG#5-HRP was used at 0.09 μg $mL^{-1}$ and Jackson IgG-HRP was used at 0.02 μg $mL^{-1}$.

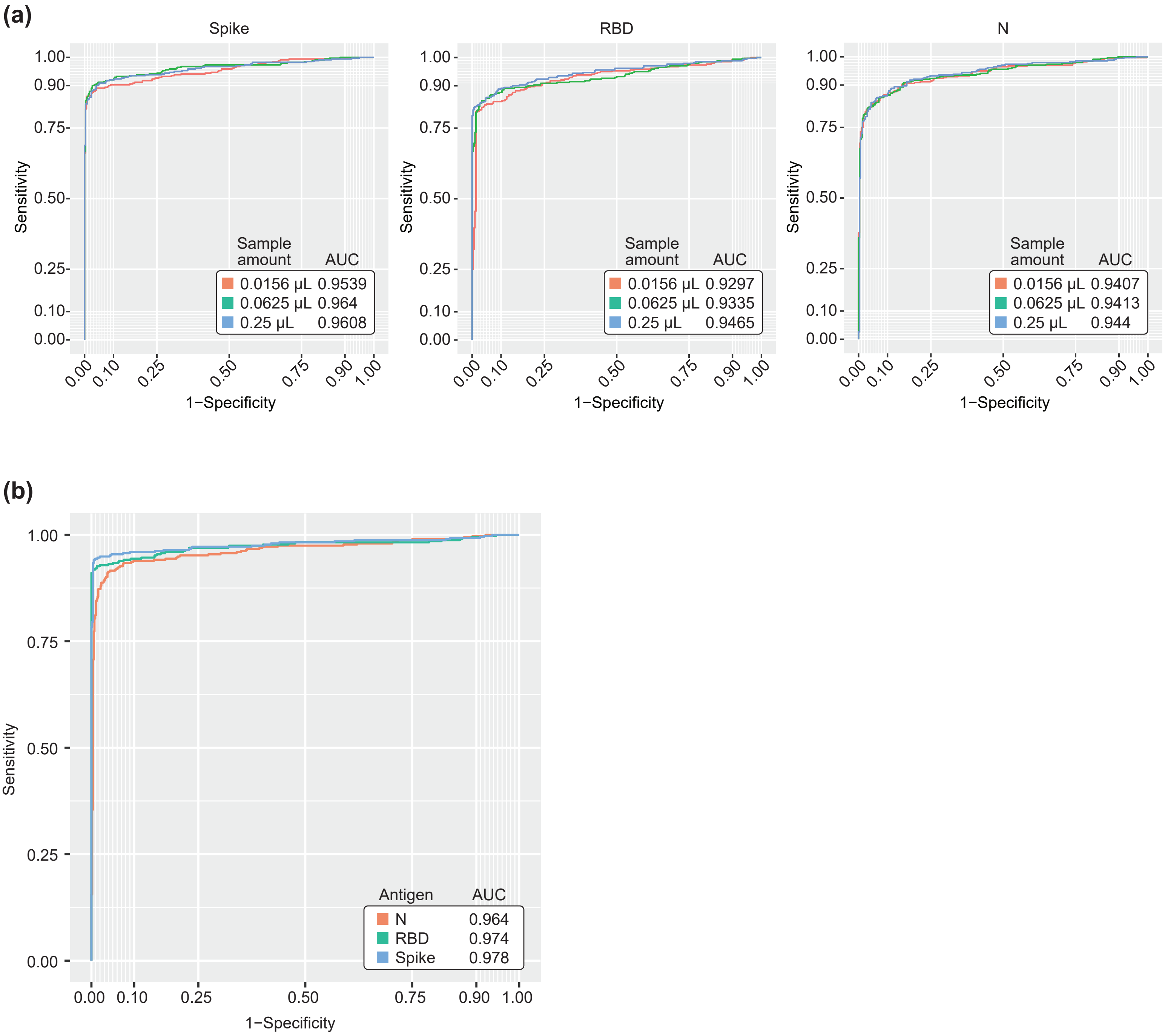


**Supplementary figure 4**. Performance of the IgG chemiluminescent ELISAs with plasma and serum. **(a)** ROC analyses of three different dilutions of negative (pre-COVID-19, *n* = 300) and positive (convalescent, *n* = 211) plasma and serum samples. The dilution is the amount of sample added per well. **(b)** ROC analysis at 0.0625 μL/well. A subset of samples from (a) (181 positive, 260 negative) were repeated and all samples from the 0.0625 μL dilution were treated as independent samples. Raw values were normalized to the 0.0156 μg mL$^{-1}$ point in the reference curve for spike and RBD and to the 0.0625 μg mL$^{-1}$ point for N. Performance statistics are shown in Table 1.

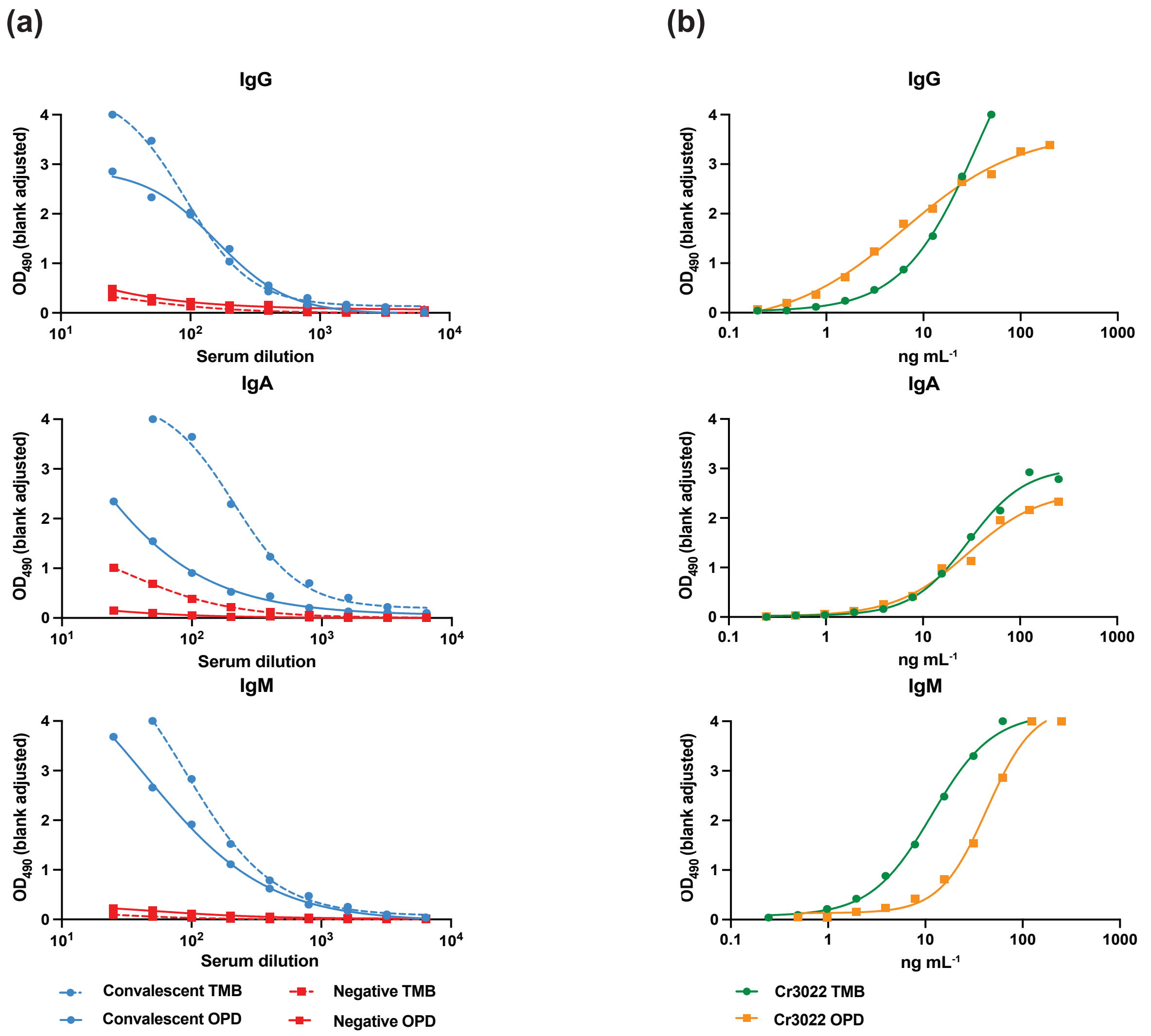


**Supplementary figure 5.** Comparison between o-Phenylenediamine dihydrochloride (OPD) and 3,3′,5,5′-tetramethylbenzidine (TMB) as colorimetric substrates for the manual 96-well plate ELISA. A final volume of 100 µL of substrate was added and incubated in the dark with shaking for 10 min before reactions were stopped by the addition of 50 µL of 3 M HCl. **(a)** A SARS-CoV-2 convalescent serum and a known negative serum were titrated to compare the relative performance of the substrates. Antibodies against the RBD (319–541, 100 ng/well) were measured using monoclonal IgG (Millipore A0170), IgA (The Jackson Laboratory 109-035-129), and IgM (The Jackson Laboratory 109-035-011) at 1:3,000. Absorbance at 490 nm was measured and blank-adjusted. **(b)** Titration of the commercial isotype-specific CR3022 clone (Absolute Antibody) against the RBD for all three main isotypes using TMB or OPD. CR3022 isotype-specific curves were generated on the same plates as the serum titration shown in (a) and were used as standard/calibration curves. TMB was more sensitive but did not substantially increase the dynamic range between the known negative and positive samples.

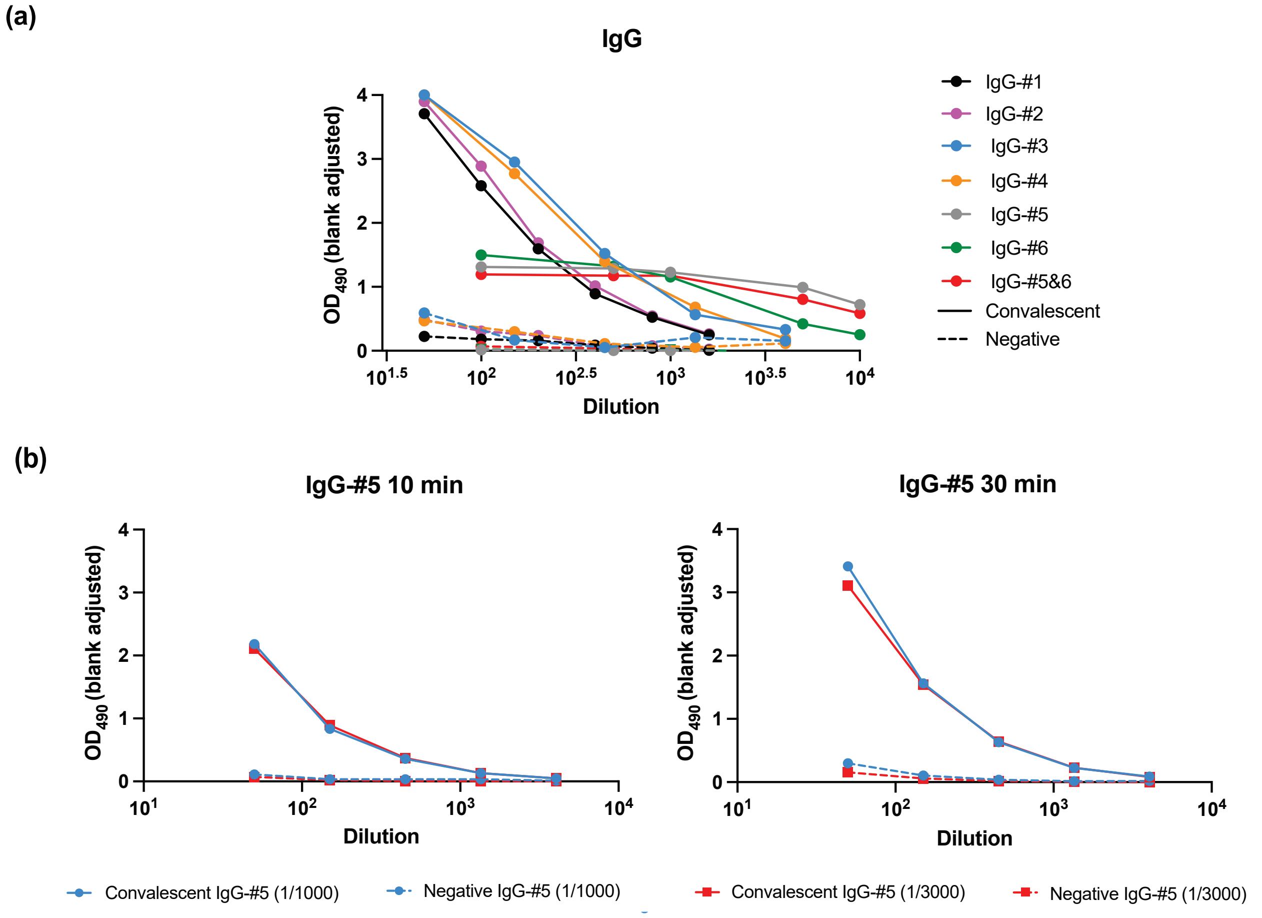


**Supplementary figure 6.** Comparison of IgG-HRP secondary antibody performance **(a)** Analysis of NRC IgG-HRP secondary (detection) antibodies. A convalescent SARS-CoV-2 serum and a known negative serum were titrated, and RBD-specific IgG antibodies were measured using different secondary antibodies at 1:3,000. Coated RBD (319–541) antigen was constant at 100 ng/well and OPD was used as the colorimetric substrate. IgG #5 and #6 are HRP fusions, while IgG-#1 to #4 are HRP conjugates that gave the highest signals. IgGs-#5 and #6 had the lowest cross-reactivity to the negative serum. Combining IgG #5 and #6 had no impact on the signal. IgG#5-HRP was chosen as the default IgG secondary for our serological assay. **(b)** Optimization of OPD development time and IgG#5-HRP concentration. Convalescent SARS-CoV-2 serum and known negative serum were titrated and RBD-specific IgG antibodies were measured using the IgG-#5-HRP at 1:1,000 and 1:3,000. Two reaction times were tested, and reactions were stopped by adding 3 M HCL. Increasing the reaction time resulted in a higher signal and increased dynamic range between the positive and negative sera. The higher concentration of IgG#5-HRP had no significant impact on the signal.

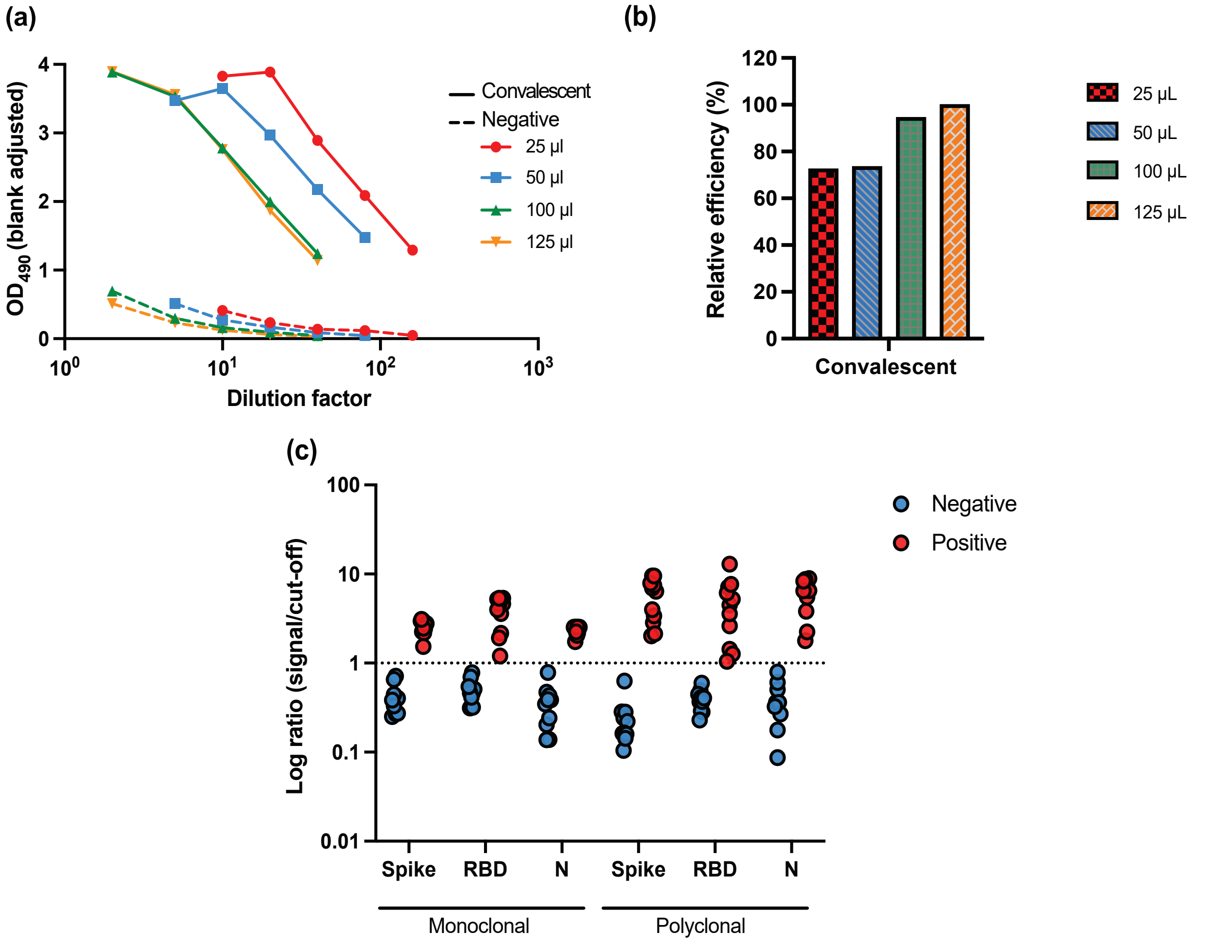


**Supplementary figure 7.** Elution and secondary antibody optimizations for IgG ELISAs using DBSs. **(a)** To determine the optimal elution volume, a DBS sample from a convalescent SARS-CoV-2 individual (CoV-19) and a known negative DBS sample (CoV-20) were eluted in four different volumes of PBS from 3 mm DBS discs at room temperature overnight with shaking in a 96-well u-bottom plate. IgG antibodies against spike (100 ng/well) were measured using polyclonal IgG-HRP (Abcam, #ab97165; at 1:3,000) as a secondary antibody and developed with OPD. **(b)** The elution efficiency was calculated from the results shown in (a). To account for the different dilutions, the IgG-CR3022 standard curve was used to calculate the titers of anti-spike IgG in each elution volume. The 125 µL elution volume had the highest titers and was set at 100%, with other conditions calculated in relation to that value. Volumes of 100 or 125 µL enabled optimal elution, and elution efficiency was significantly impacted at lower volumes. Therefore, we used 100 µL/3 mm DBS disc in our downstream serology ELISAs, mitigating the need for highly concentrated eluates while maintaining good elution efficiency. **(c)** Signal to cutoff (S/CO) ratios were calculated for a panel of 20 contrived DBS specimens of known positive and negative samples produced by the NML. The samples were tested for IgG antibodies using monoclonal (IgG-#5-HRP) and polyclonal (ab97165) secondary antibodies for spike, RBD (319–541), and N. All DBS punches were eluted in 100 µL PBS overnight at room temperature. For the monoclonal secondary the eluate was used undiluted, while for the polyclonal antibody it was diluted 1:10. The cutoff was set at two SDs of the negative distribution.

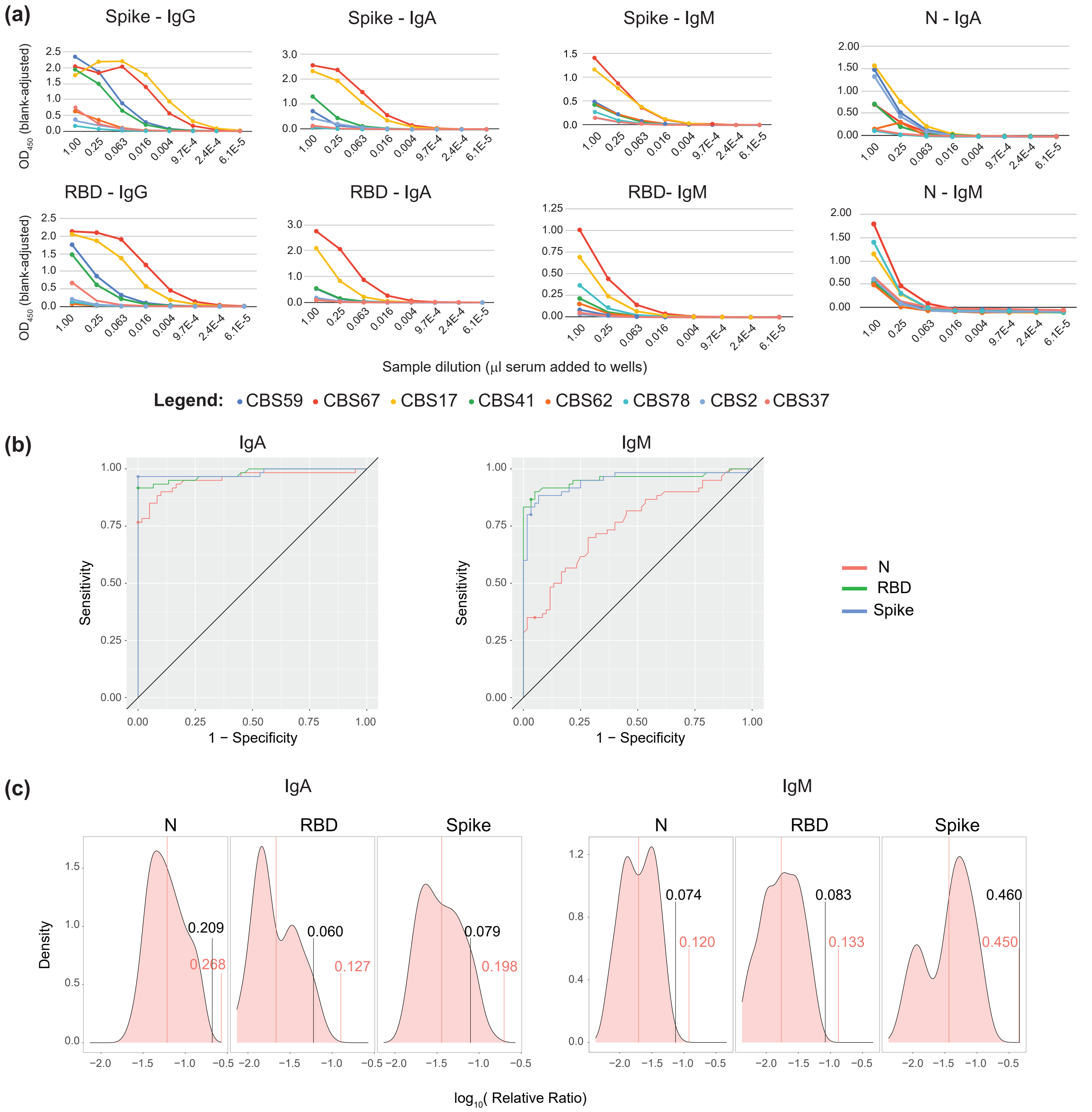


**Supplementary figure 8.** IgA and IgM ELISAs. **(a)** Titration curves of CBS samples were used to test the Jackson anti-IgA (1:10,000) and anti-IgM (1:60,000) HRP secondary antibodies using the manual assay on spike (200 ng/well), RBD (331–521, 75 ng/well), and N (25 ng/well), and to compare them against IgG curves generated using anti-IgG#5-HRP (1:10,000) for spike and RBD (see Supplementary Figure 13 for N). Due to precipitate formation in the IgA experiment at high sample concentrations, a 1:20,000 dilution was selected for subsequent experiments. **(b)** ROC analysis for IgM and IgA was performed using the automated assay on a sample set of PCR-confirmed COVID-19 cases 20–40 days post-symptom onset and negative (pre-COVID-19) samples (see Supplementary Table 4 for statistics). **(c)** Similar to IgG, we compared these thresholds to those determined by calculating three SDs from the mean of the negative control log distribution (two tests for IgA 1 month apart, *n* = 72; three tests for IgM over 2 months, *n* = 120). For IgA, this analysis suggested that the thresholds for spike and RBD based on the ROC were not sufficiently stringent. Similarly, for IgM, the thresholds for RBD and NP appeared lenient. The final thresholds selected for IgA and IgM were therefore based on three SDs from the mean of the negative controls (see Supplementary Table 3 for statistics).

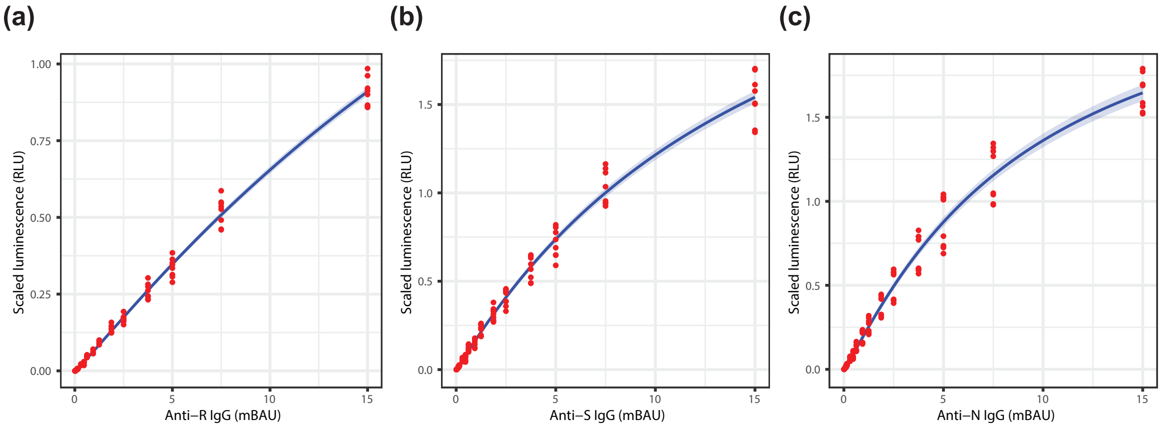


(d)

| Sample ID | Anti-R IgG | Anti-S IgG | Anti-N IgG | IgG Call |
|---|---|---|---|---|
| Sample 1 | 0.592 | 0.065 | 0.114 | Negative |
| Sample 2 | 0.237 | 0.063 | 0.108 | Negative |
| Sample 3 | 1.103 | 0.891 | 4.021 | Positive |
| Sample 4 | 0.405 | 0.078 | 0.117 | Negative |
| Sample 5 | 0.306 | 0.091 | 0.136 | Negative |
| Sample 6 | 0.269 | 0.075 | 0.099 | Negative |
| Sample 7 | 0.313 | 0.111 | 0.150 | Negative |
| Sample 8 | 12.247 | 7.179 | 23.931 | Positive |
| Sample 9 | 0.198 | 0.099 | 0.154 | Negative |
| Sample 10 | 0.236 | 0.089 | 0.145 | Negative |
| Sample 11 | 8.017 | 6.075 | 30.774 | Positive |
| Sample 12 | 10.387 | 7.173 | 12.667 | Positive |
| Sample 13 | 0.547 | 0.134 | 5.360 | Negative |
| Sample 14 | 9.428 | 6.854 | 13.167 | Positive |
| Sample 15 | 0.369 | 0.137 | 0.210 | Negative |
| Sample 16 | 11.869 | 6.277 | 79.842 | Positive |
| Sample 17 | 0.354 | 0.127 | 0.142 | Negative |
| Sample 18 | 0.292 | 0.129 | 0.131 | Negative |
| Sample 19 | 2.320 | 2.819 | 1.185 | Positive |
| Sample 20 | 0.294 | 0.104 | 0.129 | Negative |
| Sample 21 | 0.137 | 0.058 | 0.060 | Negative |
| Sample 22 | 3.462 | 5.861 | 23.680 | Positive |
| Sample 23 | 1.815 | 1.512 | 0.493 | Positive |
| Sample 24 | 0.337 | 0.108 | 0.184 | Negative |
| Sample 25 | 0.657 | 0.137 | 0.215 | Negative |
| Sample 26 | 0.205 | 0.067 | 0.118 | Negative |

**Supplementary figure 9**. Conversion of serology data to WHO BAUs and validation of SARS-CoV-2 IgG profiling performed on the uOttawa automated serology platform. Calibrants used were the National Institute for Biological Standards and Control (NIBSC) reference panel (code: 20/268) and the First WHO International Standard (20/136). **(a)–(c)** Titration of the NIBSC 20/136 pooled convalescent serum for IgG using spike, RBD, and N. Linear regression using scaled luminescence values (as detailed in Supplementary Figure 11) and the viral titre in BAU μL$^{-1}$ is shown. **(d)** Titers (BAU mL$^{-1}$) and calls for the set of representative samples analyzed in Supplementary figure 11. Yellow, green, and red indicate samples above the 3% FDR threshold, overall negative samples, and overall positive samples, respectively. Samples were considered positive for SARS-CoV-2 IgG when both anti-S and anti-N IgG were detected above their thresholds.

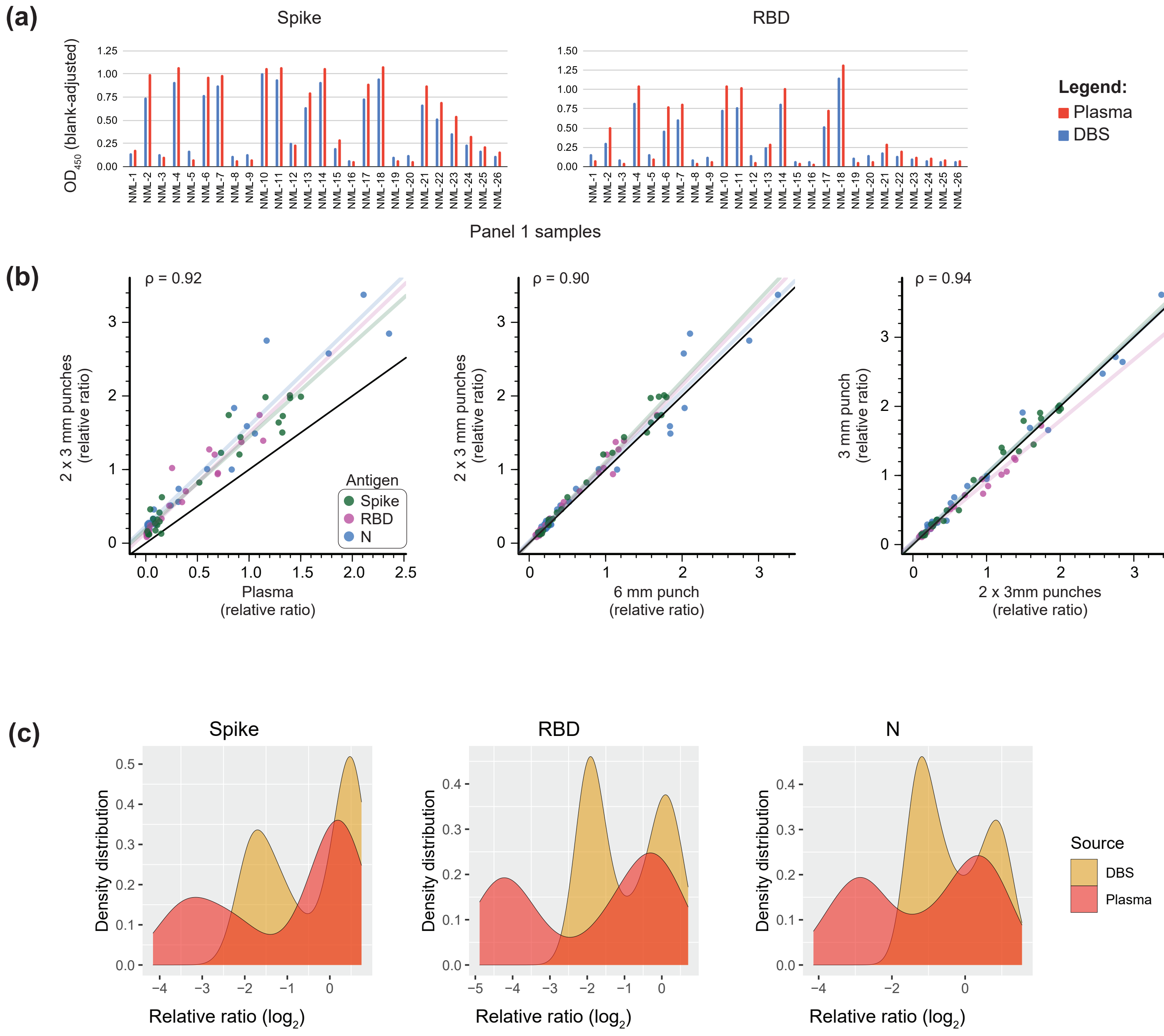


**Supplementary figure 10.** DBS Optimization. **(a)** 6 mm punches were taken from DBSs and eluted in 250 µL PBST plus 1% Triton X-100 (a 500 µL elution volume was initially tested but was too dilute). 50 µL of eluate or 1 µL of plasma diluted in 50 µL 1% w/v milk power in PBST were added to wells coated with 200 ng spike or 75 ng RBD (Rini) for colorimetric ELISA using 1:60,000 Jackson anti-IgG HRP as the secondary antibody. Blank-subtracted samples were normalized to positive serum pools analyzed on the same plate. **(b)** Correlation of DBSs to plasma (left panel) and different size DBS punches to each other (middle and right panels) for the indicated antigen (*n* = 26, NML panel 3, 1:4 dilution of DBS eluate). **(c)** Density distributions of DBS and matching plasma samples from NML panels 1–3 (*n* = 96). The shift in the DBS density distribution means that thresholds established for plasma cannot be used directly to call positives using DBSs.

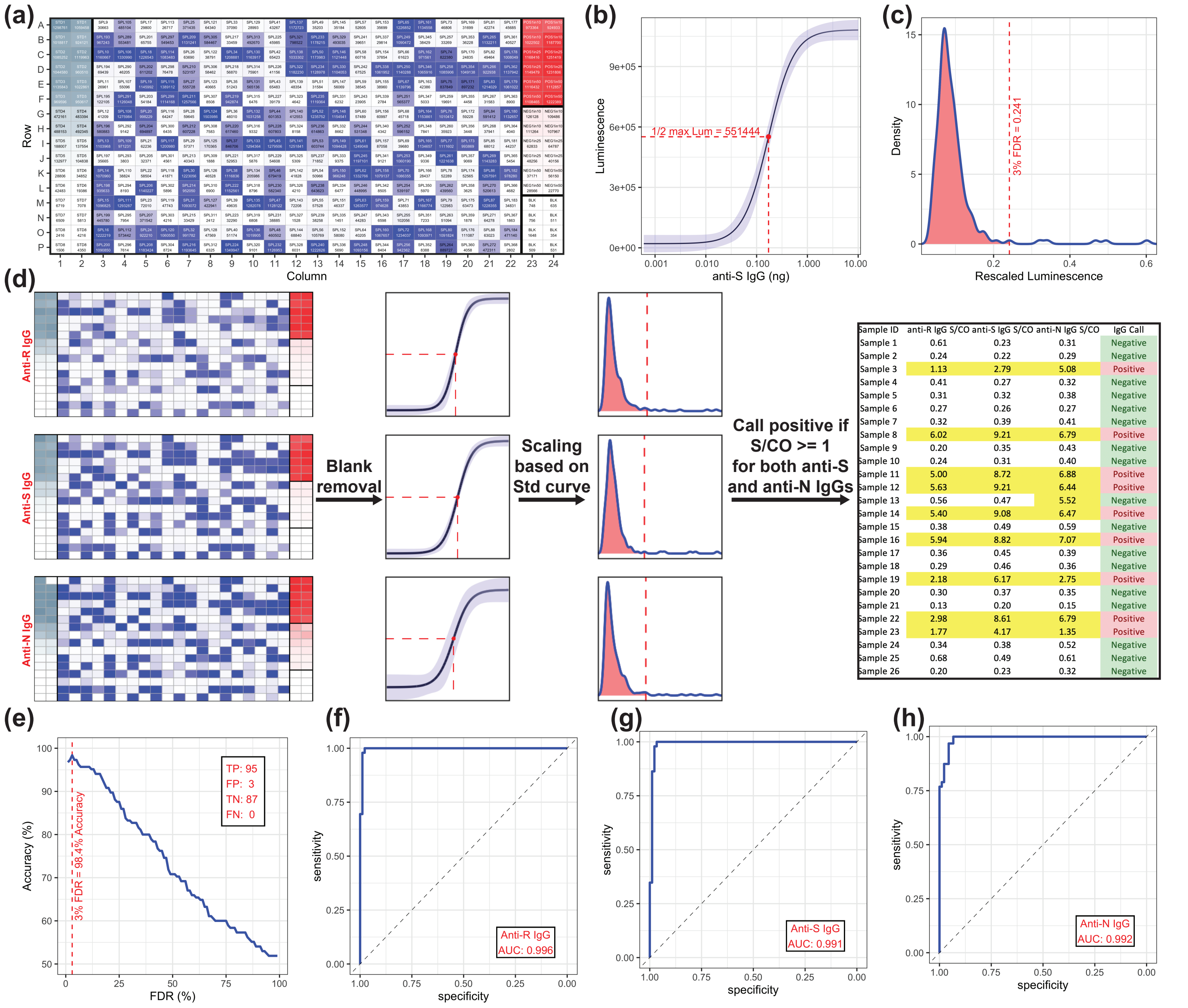


**Supplementary figure 11.** Processing and validation of the Ottawa automated serology platform. **(a)** Raw luminescence values generated by the automated platform, shown on the established plate map. Color intensities are relative to luminescence values from a prototypical run. Standards, test samples, and controls are colored gray, blue, and red respectively. For each well, sample identifications and luminescence values are also indicated. **(b)** The IC50 luminescence value of the standard curve is used to scale sample values. The curve is derived from the standard samples shown in (a) and modelled using a four-parameter log-logistic function. The shaded blue area indicates the 95% confidence interval, and both the red point and dashed lines specify the inflection point of the curve. The luminescence value at the inflection point is used to scale all the values for a given antigen on the plate for further processing. **(c)** FDR determination using pre-pandemic samples. Anti-S IgG luminescence values from DBSs were rescaled as in (b), and a density distribution was used to determine a threshold corresponding to a 3% FDR (dashed line). The red area includes 97% of the scaled luminescence values. **(d)** Pipeline used to call SARS-CoV-2 IgG positive samples. For each plate, blank values were subtracted, the luminescence was rescaled based on luminescence values at the inflection points of the respective standard curves, and signal to cutoff (S/CO) values were calculated. Samples were considered overall positive for SARS-CoV-2 IgG when both anti-S and anti-N IgG were detected with S/CO ≥ 1. Yellow, green, and red backgrounds in the table on the right indicate S/CO ≥ 1, negative, and positive samples, respectively. **(e)** Determination of the optimal FDR threshold for SARS-CoV-2 IgG positive calls. FDRs of 1–100% were plotted as in C for anti-RBD, -spike, and -N IgG. For each FDR value, the number of true positive (TP), false positive (FP), true negative (TN), and false negative (FN) samples were calculated based on DBS samples provided by the NML (Panel 4) to determine the calling accuracy (i.e. the percentage of TP+TN samples). A maximum accuracy of 98.4% was obtained with a 3% FDR. The inset shows the sample distributions for the four specificity statistics at a 3% FDR. **(f–h)** ROC analyses of the three IgGs used in the analysis. S/CO values and true sample identifications from the NML were used as parameters for each analysis. Calculated AUC values are indicated.

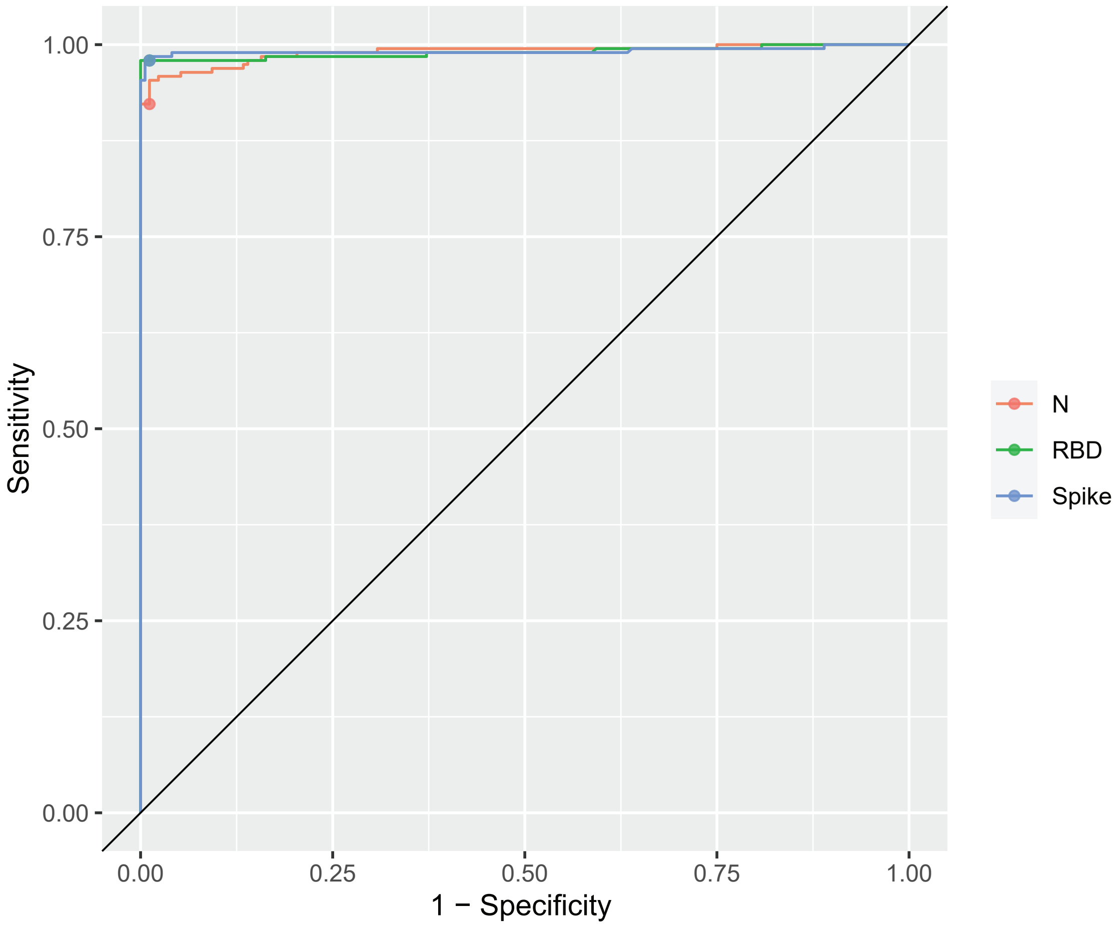


**Supplementary figure 12.** ROC curve for NML panel 4 DBS IgG analysis. DBS eluates were analyzed for antibodies against the three antigens as indicated. Samples were diluted 1:4 and analyzed on the F7 platform in Toronto in 384-well chemiluminescent format. Statistics and sample numbers are provided in Table 3.

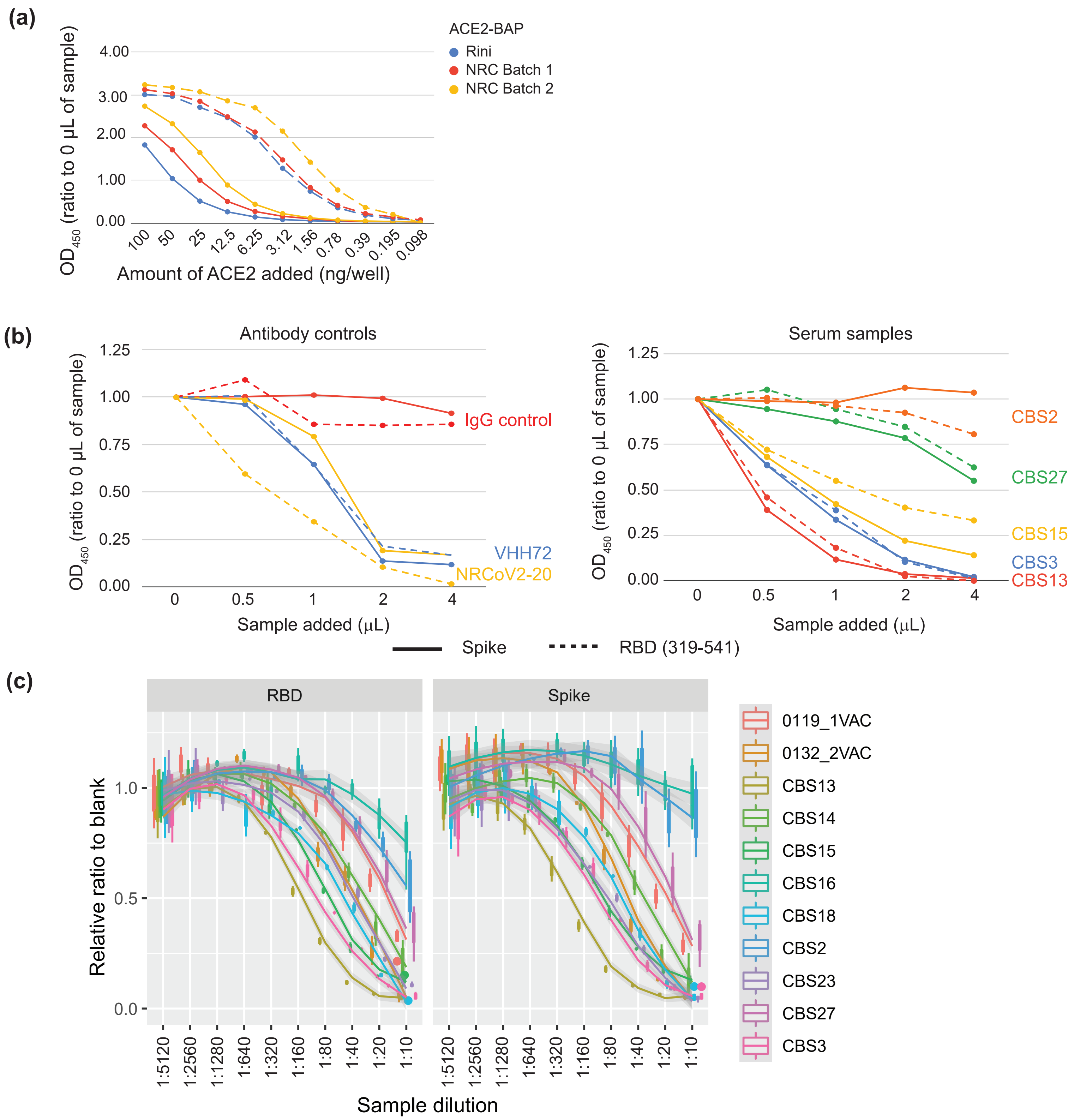


**Supplementary figure 13.** Optimization and testing of NRC reagents for snELISA. **(a)** To determine the amount of ACE2-BAP to add to the manual snELISA, 500 ng of VHH72-Fc (positive control, solid line) or IgG (negative control, dashed line) was added to a plate coated with 200 ng/well of spike. The indicated amount of ACE2 from the NRC was compared to the validated ACE2 reagent from the Rini lab. Batch 2 ACE2 was enriched for biotinylation using a Monomeric Avidin Ultralink column (Pierce). For the colorimetric assay, 6.25 ng/well was selected as the optimal concentration for ACE2-BAP as it was most comparable to biotinylated ACE2 from the Rini lab, showed good separation between the controls, and maintained a high signal for the IgG control. **(b)** Using the optimized amount of ACE2-BAP Batch 1 (6.25 ng/well for spike, 1.56 ng/well for RBD), titration curves were generated for spike and its RBD using antibody controls (left panel) and CBS samples (right panel). Spike and RBD had similar profiles, with clear separation between controls and samples. **(c)** Dose-response curves for spike and RBD, linked to Figure 4A. Values are normalized to the blank (no sample added).

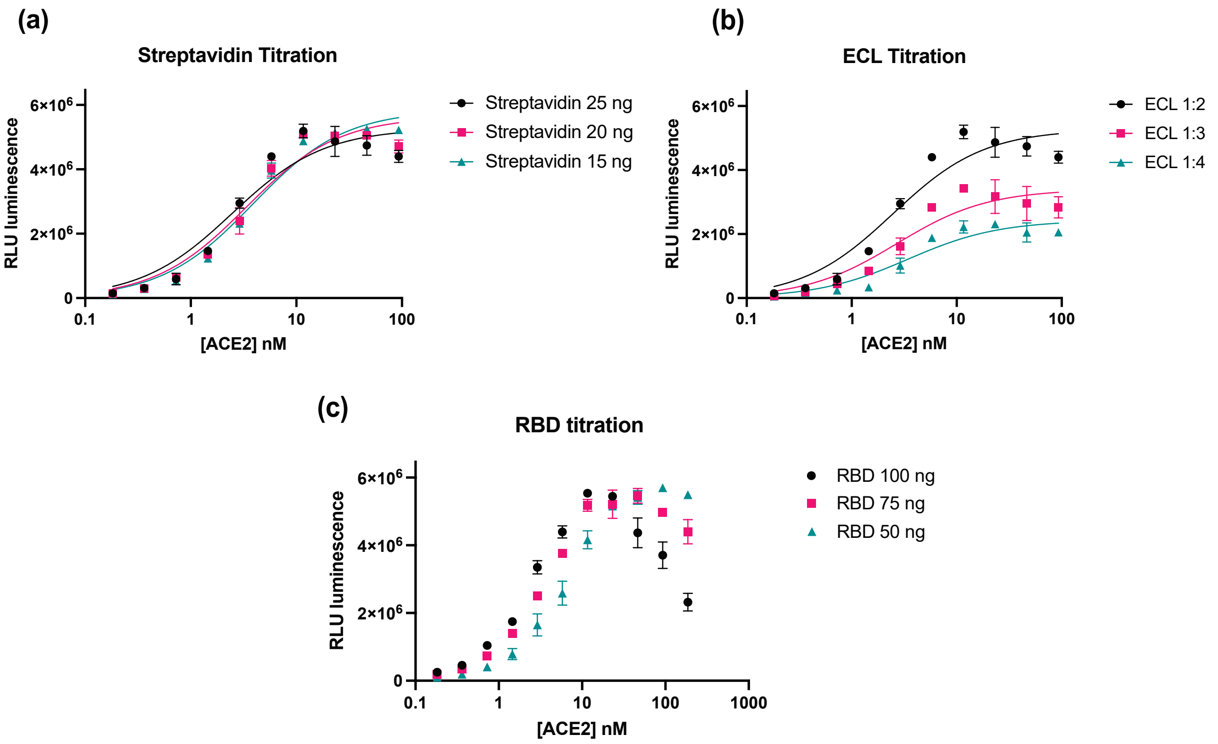


**Supplementary figure 14.** Optimization of the snELISA for the automated platform. **(a)** The streptavidin-HRP polymer (Sigma #S2438) was titrated (25, 20, and 15 ng/well) in 100 ng/well RBD and a substrate to sample ratio of 1:2 while varying the ACE2 concentration from 0.25 to 128 ng/well. Adjusting the streptavidin-HRP concentration had limited impact, highlighting its saturation. **(b)** The luminescence substrate (Thermo Fisher Scientific, #37069) was titrated by dilution in MilliQ $H_2O$ (1:2, 1:3, and 1:4) while maintaining the RBD at 100 ng/well and the streptavidin-HRP at 25 ng/well and varying the ACE2 concentration as in (a). **(c)** The coated RBD concentration was titrated (100, 75, and 50 ng/well) while maintaining streptavidin-HRP at 25 ng/well and the luminescence substrate at 1:2 and varying the ACE2 concentration from 0.25 to 256 ng per well. A profound hook-effect was noticed at high ACE2 concentrations.

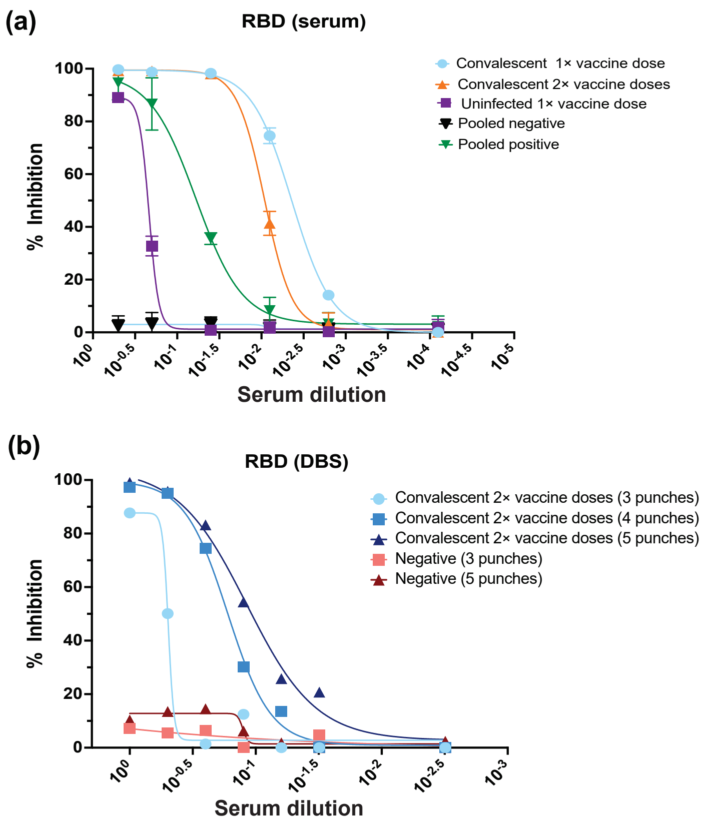


**Supplementary figure 15.** Optimized snELISA for the automated platform. **(a)** Inhibition of ACE2 binding to RBD (319–541) in the presence of different sources of serum. A 6-point titration of the serum enables the calculation of EC50 values. Serum samples were from a convalescent SARS-CoV-2 individual 3 weeks-post first vaccination (Comirnaty) and 3 weeks post-second vaccination (Comirnaty), an individual with no prior SARS-CoV-2 infection (Surveillance) 3 weeks post-first vaccination (Comirnaty), and pooled sera from individuals ($n$ = 100) with or without prior SARS-CoV-2 infection. The assay was performed with 100 ng/well of RBD (319–541), challenged with 6.5 ng/well of ACE2, and detected with 25 ng/well of streptavidin-HRP polymer. **(b)** For DBS neutralization, a comparison between three, four, and five (3 mm) punches eluted in 100 µL was performed on DBS samples from the twice-vaccinated convalescent individual in (a) and a known negative sample. Four and five punches had similar performance, which was significantly lower with three. To calculate the percent inhibition, luminescence values were scaled to assays without serum (max signal). Serum samples were analyzed in quadruplicate while DBS samples were analyzed once.

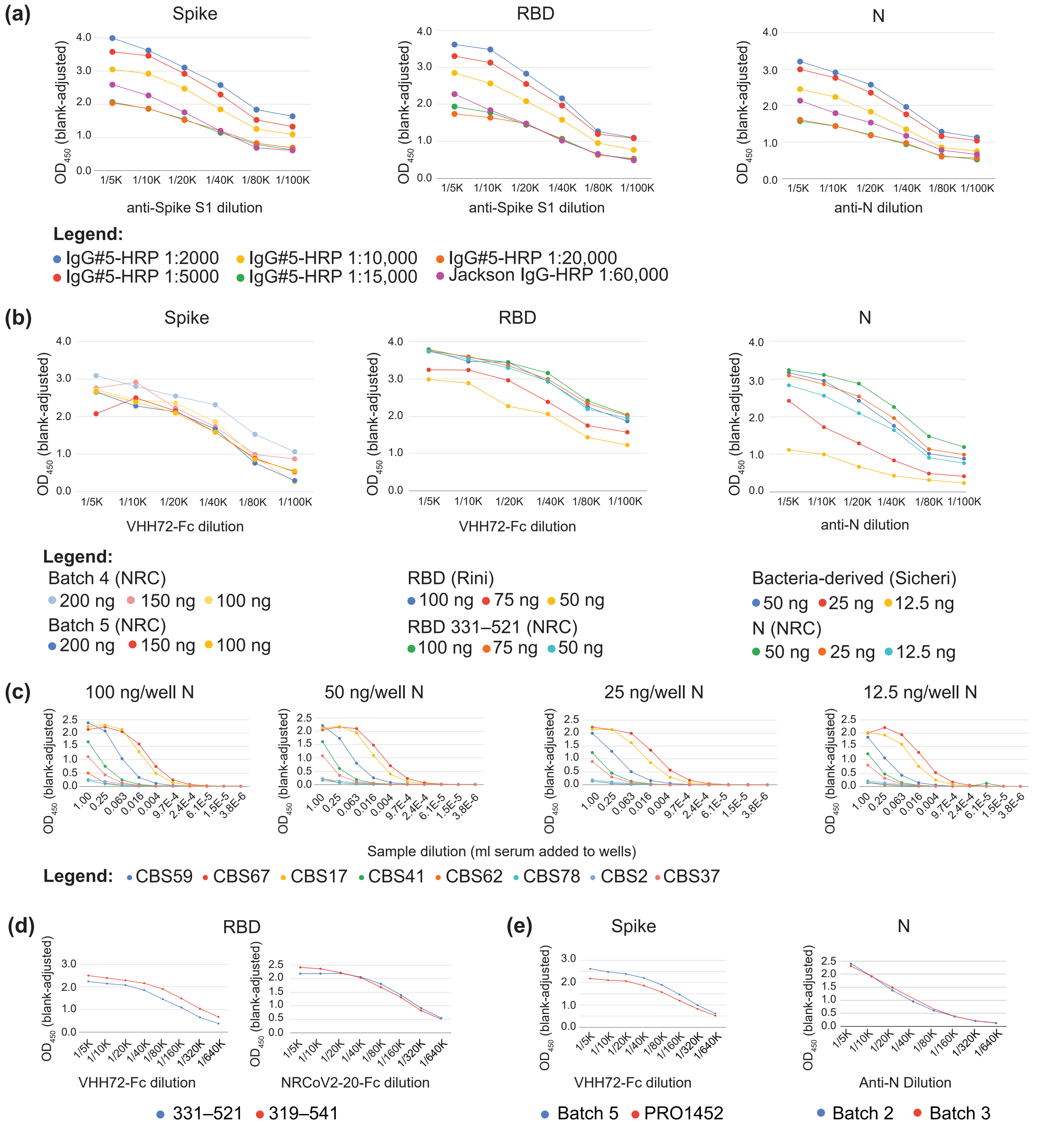


**Supplementary figure 16.** Optimization of the NRC reagents. **(a)** Different dilutions of IgG#5-HRP were compared to Jackson IgG-HRP by colorimetric ELISA. Antigen amounts: spike (200 ng/well, NRC), RBD (75 ng/well, Rini), and N (25 ng/well, Sicheri). Anti-spike S1 IgG (GenScript, 1 mg mL$^{-1}$ stock) or anti-N IgG (GenScript, 1 mg mL$^{-1}$ stock) were diluted as indicated. Based on the similarity of its curve with that of the Jackson antibody, the 1:10,000 dilution of IgG#5-HRP was selected. **(b)** Different dilutions of NRC antigens were compared to previously used antigens [8]. VHH72-Fc and anti-N IgG antibodies were diluted as indicated. Based on the similarity of the curves to those of prior antigens, the same were selected for spike (200 ng/well) and its RBD (75 ng/well). **(c)** Due to the difference in signal between N produced in bacteria vs. in mammalian cells, we performed an extended titration analysis of CBS samples to determine the amount of antigen. Based on the shape of the curves and to conserve reagents, 25 ng/well of N was chosen for manual colorimetric assays. **(d)** Comparison of two different RBD proteins (75 ng/well). **(e)** Comparison of two different batches of spike SmT1 (200 ng/well) and N (25 ng/well). For B to E, IgG#5-HRP was used at 1:10,000.

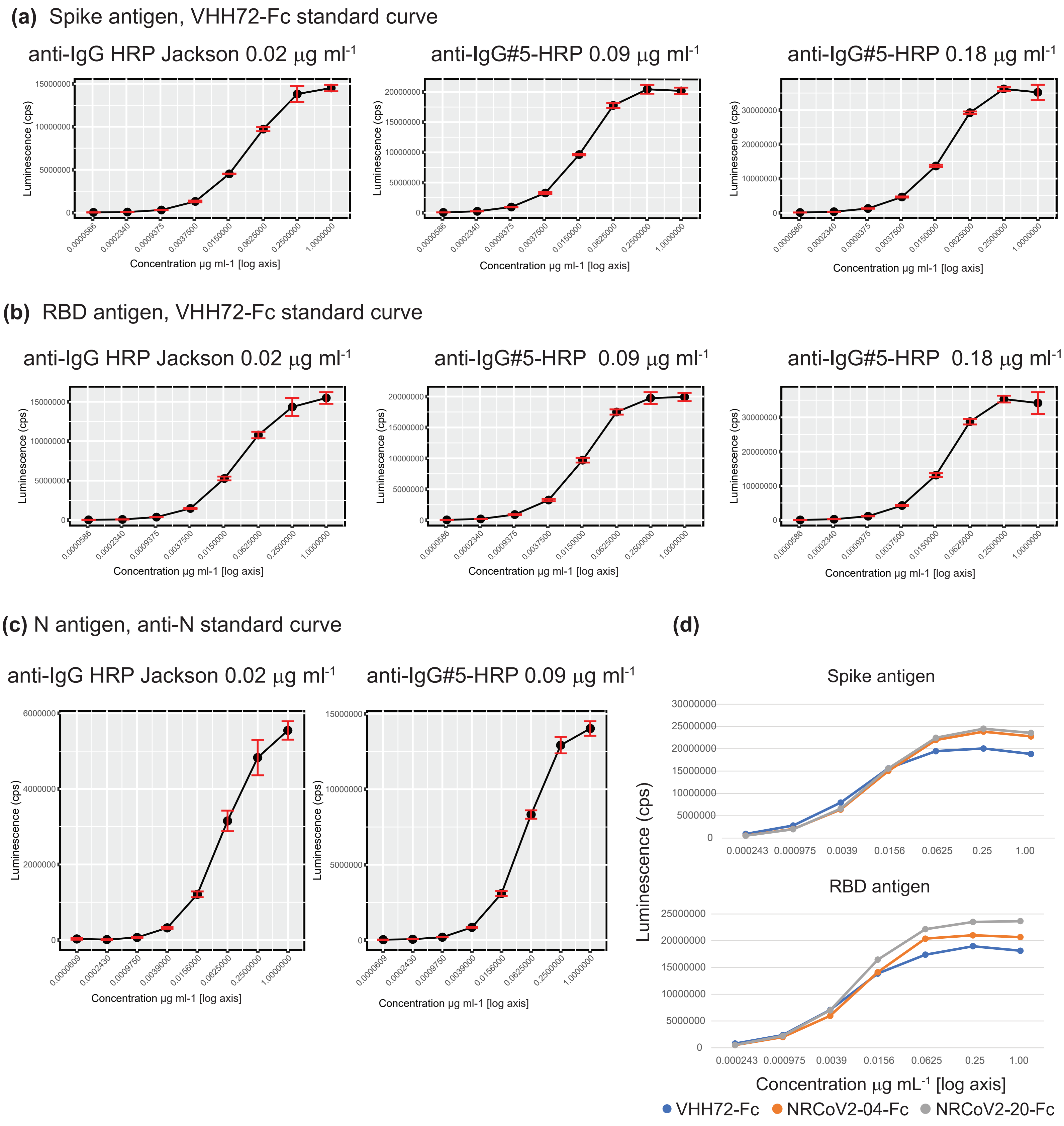


**Supplementary figure 17.** Optimization of IgG#5-HRP and recombinant antibodies for chemiluminescent assays. **(a)** and **(b)** Two different concentrations of IgG#5-HRP (selected based on the colorimetric assay) were compared to the Jackson IgG secondary antibody. Note that the scales differ between graphs to illustrate their similar binding curves. For spike and its RBD, 0.09 and 0.18 μg $mL^{-1}$ produced similar curve profiles to the Jackson secondary, with all curves beginning to plateau at 0.25 μg $mL^{-1}$. Increased secondary antibody did result in higher luminescence signal, suggesting that the antigen is the limiting factor at higher primary antibody levels. For subsequent experiments, 0.09 μg $mL^{-1}$ was selected. **(c)** For N, 0.09 μg $mL^{-1}$ of IgG#5-HRP was compared to the Jackson secondary. **(d)** Two alternative recombinant antibodies against the RBD were tested by ELISA for use in standard curves with the spike and RBD antigens. Both alternates had similar binding profiles to VHH72. Antigens for all tests: spike (batch 5, 50 ng/well), RBD (331–521, 20 ng/well), N (7 ng/well). Antigen concentrations for automated chemiluminescent assays were decreased 3.5–4-fold from the optimal concentrations for the manual colorimetric assay.
