## Supplementary Tables 1-4 for "A scalable serology solution for profiling humoral immune responses to SARS-CoV-2 infection and vaccination"

**Supplementary table 1.** Reagent information.

| **Protein** | **Protein type** | **Expression mode** | **Typical purified yield (mg L$^{-1}$)** | **% Main peak purity by SEC** |
|---|---|---|---|---|
| spike (SMT1) | 570 kDa trimer | CHO stable pool | 370 | 97 |
| RBD (331–521) | 37 kDa monomer | CHO transient | 25 | 98 |
| RBD (319–541) | 37 kDa monomer | CHO transient | 30 | 96 |
| nucleocapsid (NCAP) | 300 kDa hexamer[†] | CHO transient | 60 | 99 |
| VHH72-hFc1X7 | 100 kDa dimer | CHO transient | 250 | 98 |
| NRCoV2-04-hFc1X7 | 90 kDa dimer | CHO transient | 100 | 91 |
| NRCoV2-20-hFc1X7 | 100 kDa dimer | CHO transient | 250 | 98 |
| IgG#5-HRP | 270 kDa mAb | CHO transient | 15 | 60 |
| ACE2-BAP | 90 kDa monomer | CHO transient | 150 | 99 |

[†]Hexamer predicted based on HPLC-SEC elution volume.

**Supplementary table 2.** Comparison of manual and automated ELISAs.

| Reagent | Manual ELISA (96-well colorimetric) | | Automated ELISA (384-well chemiluminescent) | |
|---|---|---|---|---|
| | **Toronto** | **Ottawa** | **Toronto** | **Ottawa** |
| **Antibody detection ELISA** | | | | |
| spike | 200 ng/well | 100 ng/well | 50 ng/well | 50 ng/well |
| RBD (331–521) | 75 ng/well | N/A | 20 ng/well | N/A |
| RBD (319–541) | 75 ng/well | 100 ng/well | 20 ng/well | 50 ng/well |
| N | 25 ng/well | 100 ng/well | 7 ng/well | 50 ng/well |
| spike/RBD control antibody (Toronto: VHH72-hFc1X7 for IgG, hIGM2001 to IgM, CR3022 for IgA, Ottawa: CR3022) | 8-point curve of 2-fold dilutions starting at 10 ng/well | 10-point curve of 2-fold dilutions starting at 25 ng/well for IgM/IgA and 20 ng/well for IgG | 8-point curve of 4-fold dilutions starting at 10 ng/well for IgG/IgA; 20 ng/well, 10 ng, then an 8-point curve of 4-fold dilutions for IgM | 8-point curve of 2-fold dilutions starting at 5 ng/well, 4-fold dilutions starting at 0.039 ng/well |
| N control antibody (HC2003 for IgG, CR3018 for IgM and IgA) | 8-point curve of 2-fold dilutions starting at 10 ng/well | N/A | 20 ng/well, 10 ng/well, then an 8-point curve of 4-fold dilutions | 8-point curve of 2-fold dilutions starting at 5 ng/well, 4-fold dilutions starting at 0.039 ng/well |
| IgG#5-HRP | 3 ng/well (1:10K) | 10 ng/well (1:3K) | 0.9 ng/well (1:6.7K) | 1.1 ng/well (1:5.4K) |
| Anti-IgA-HRP | 2 ng/well (1:20K) | 13 ng/well (1:3K) | 0.8 ng/well (1:10K) | 1 ng/well (1:8K) |
| Anti-IgM-HRP | 0.67 ng/well (1:60K) | 13 ng/well (1:3K) | 0.66 ng/well (1:12K) | 0.83 ng/well (1:9.6K) |
| Substrate | 50 µL TMB/50 µL sulphuric acid | 100 µL OPD/50 µL 3 M hydrochloric acid | 1:4 dilution of ELISA Pico | 1:2 dilution of ELISA Pico |
| **snELISA** | | | | |
| spike | 200 ng/well | N/A | 50 ng/well | 100 ng/well |
| RBD (319–541) | 100 ng/well | N/A | 34 ng/well | 100 ng/well |
| ACE2-BAP | 6.25 ng/well | N/A | 2.08 ng/well (NRC), 17 ng/well (Rini) | 6.5 ng/well |
| Streptavidin-HRP | 25 (1:2K) ng/well | N/A | 15 ng/well | 25 ng/well |
| Substrate | 50 µL TMB/50 µL sulphuric acid | N/A | 1:4 dilution of ELISA Pico | 1:2 dilution of ELISA Pico |

**Supplementary table 3**. ELISA performance statistics for plasma or serum IgA and IgM (Toronto).

| | **spike** | | **RBD** | | **N** | | **≥ 2 positive antigens** | |
|---|---|---|---|---|---|---|---|---|
| | **IgA** | **IgM** | **IgA** | **IgM** | **IgA** | **IgM** | **IgA** | **IgM** |
| **ROC analysis** | | | | | | | | |
| **AUC** | 0.982 | 0.949 | 0.978 | 0.955 | 0.954 | 0.759 | N/A | N/A |
| **Cut point** | 0.079 | 0.460 | 0.060 | 0.083 | 0.209 | 0.074 | N/A | N/A |
| **FPR** | 0.017 | 0.017 | 0.017 | 0.017 | 0.017 | 0.017 | 0.000 | 0.000 |
| **TPR** | 0. 967 | 0.800 | 0.917 | 0.833 | 0.767 | 0.333 | 0.917 | 0.767 |
| **3 SDs from the mean of the negative controls** | | | | | | | | |
| **Cut point** | 0.198 | 0.450 | 0.127 | 0.133 | 0.268 | 0.120 | N/A | N/A |
| **FPR** | 0.000 | 0.017 | 0.000 | 0.000 | 0.000 | 0.000 | 0.000 | 0.000 |
| **TPR** | 0.933 | 0.800 | 0.833 | 0.733 | 0.750 | 0.217 | 0.883 | 0.733 |
| **Samples used to assess performance**[†] | | | | | | | | |
| Patients with COVID-19 20–40 d post-symptom onset | | 60 | | | | | | |
| Pre-COVID-19 (negative) controls | | 60 | | | | | | |

[†]NML panel 4. Each sample was analyzed in duplicate (the unique numbers of samples are shown). Replicates were treated as separate samples in the ROC analysis. Negative samples were excluded from the analysis if the replicates were > 4 SDs from the mean.

**Supplementary table 4.** Conversion of relative ratios to international units (BAU $mL^{-1}$) for plasma or serum.

<table>
<tr><td colspan="7">Nucleocapsid†</td></tr>
<tr><td colspan="2">Dilution fold (d)</td><td colspan="2">160</td><td colspan="2">2560</td><td rowspan="3">Note</td></tr>
<tr><td colspan="2">Volume equivalent (μL)</td><td colspan="2">0.0625</td><td colspan="2">0.00391</td></tr>
<tr><td>Relative Ratio (RR)</td><td>$log_2$(RR)</td><td>$log_2$(BAU $mL^{-1}$)</td><td>BAU $mL^{-1}$</td><td>$log_2$(BAU $mL^{-1}$)</td><td>BAU $mL^{-1}$</td></tr>
<tr><td>2</td><td>1.0</td><td>8.38</td><td>333.98</td><td>12.38</td><td>5,343.76</td><td>Upper limit of linear range</td></tr>
<tr><td>1</td><td>0.0</td><td>6.98</td><td>126.34</td><td>10.98</td><td>2,021.37</td><td></td></tr>
<tr><td>0.5</td><td>-1.0</td><td>5.58</td><td>47.79</td><td>9.58</td><td>764.62</td><td></td></tr>
<tr><td>0.396</td><td>-1.3</td><td>5.11</td><td>34.46</td><td>9.16</td><td>571.19</td><td>Positivity Threshold</td></tr>
<tr><td>0.25</td><td>-2.0</td><td>4.18</td><td>18.08</td><td>8.17</td><td>289.23</td><td></td></tr>
<tr><td>0.125</td><td>-3.0</td><td>2.77</td><td>6.84</td><td>6.77</td><td>109.41</td><td></td></tr>
<tr><td>0.0625</td><td>-4.0</td><td>1.37</td><td>2.59</td><td>5.37</td><td>41.38</td><td>Lower Limit of linear range</td></tr>
<tr><td colspan="7">†Formula: $log_2$(BAU $mL^{-1}$ @ sample dilution fold d) = ($log_2$(RR) - 0.243)/ 0.713 + $log_2$(d). See Figure 3D for a graph plotting the international standard curve used to calculate the conversion formula and its relationship to the reference curve. With standard 1:160 and 1:2560 dilutions, the range is 2.59 to 5343.8 BAU $mL^{-1}$.</td></tr>
<tr><td colspan="7">RBD‡</td></tr>
<tr><td colspan="2">Dilution fold (d)</td><td colspan="2">160</td><td colspan="2">2560</td><td rowspan="3">Note</td></tr>
<tr><td colspan="2">Volume equivalent (μL)</td><td colspan="2">0.0625</td><td colspan="2">0.00391</td></tr>
<tr><td>Relative Ratio (RR)</td><td>$log_2$(RR)</td><td>$log_2$(BAU $mL^{-1}$)</td><td>BAU $mL^{-1}$</td><td>$log_2$(BAU $mL^{-1}$)</td><td>BAU $mL^{-1}$</td></tr>
<tr><td>1</td><td>0.0</td><td>8.12</td><td>278.37</td><td>12.12</td><td>4,453.99</td><td>Upper limit of linear range</td></tr>
<tr><td>0.5</td><td>-1.0</td><td>6.82</td><td>112.63</td><td>10.81</td><td>1,802.02</td><td></td></tr>
<tr><td>0.25</td><td>-2.0</td><td>5.51</td><td>45.57</td><td>9.51</td><td>729.07</td><td></td></tr>
<tr><td>0.186</td><td>-2.4</td><td>4.95</td><td>30.97</td><td>8.95</td><td>495.58</td><td>Positivity Threshold</td></tr>
<tr><td>0.125</td><td>-3.0</td><td>4.20</td><td>18.44</td><td>8.20</td><td>294.97</td><td></td></tr>
<tr><td>0.0625</td><td>-4.0</td><td>2.90</td><td>7.46</td><td>6.90</td><td>119.34</td><td></td></tr>
<tr><td>0.03125</td><td>-5.0</td><td>1.59</td><td>3.02</td><td>5.59</td><td>48.28</td><td>Lower Limit of linear range</td></tr>
<tr><td colspan="7">‡Formula: $log_2$(BAU $mL^{-1}$ @ sample dilution fold d) = ($log_2$(RR) + 0.612)/ 0.766 + $log_2$(d). With standard 1:160 and 1:2560 dilutions, the range is 3.02 to 4,449.72 BAU $mL^{-1}$.</td></tr>
</table>

| Spike[§] | | | | | | |
|---|---|---|---|---|---|---|
| Dilution fold (d) | | 160 | | 2560 | | Note |
| Volume equivalent (μL) | | 0.0625 | | 0.00391 | | |
| Relative Ratio (RR) | $log_2$(RR) | $log_2$(BAU $mL^{-1}$) | BAU $mL^{-1}$ | $log_2$(BAU $mL^{-1}$) | BAU $mL^{-1}$ | |
| 1 | 0.0 | 6.55 | 93.80 | 10.55 | 1,500.80 | Upper limit of linear range |
| 0.5 | -1.0 | 5.28 | 38.75 | 9.27 | 619.95 | |
| 0.25 | -2.0 | 4.00 | 16.01 | 8.00 | 256.09 | |
| 0.19 | -2.4 | 3.50 | 11.28 | 7.49 | 180.45 | Positivity Threshold |
| 0.125 | -3.0 | 2.72 | 6.61 | 6.72 | 105.78 | |
| 0.0625 | -4.0 | 1.45 | 2.73 | 5.45 | 43.70 | |
| 0.03125 | -5.0 | 0.17 | 1.13 | 4.17 | 18.05 | Lower Limit of linear range |

[§]Formula: $\log_2(\text{BAU mL}^{-1} \text{ @ sample dilution fold d}) = (\log_2(\text{RR}) - 0.604)/0.784 + \log_2(d)$. With standard 1:160 and 1:2560 dilutions, the range is 1.13 to 1,500.80 BAU $mL^{-1}$.
